# Validation of the Comprehensive Online Sleep Monitoring Scale (COSMOS) in a Large Population Sample

**DOI:** 10.1101/2024.10.21.24315765

**Authors:** Laila Rida, Konstantinos Ioannidis, Samuel R. Chamberlain, Jon E. Grant, Peter Hellyer, Valentina Giunchiglia, William Trender, Karla V. Allebrandt, Sukhi Shergill, Steven CR Williams, Adam Hampshire

## Abstract

Researchers are increasingly studying cognitive and psychological constructs using automated online tools due to advantages in scalability, repeatability, accessibility and affordability. The online assessment of sleep presents a challenge, as the most popular instruments for reporting different aspects of sleep were originally designed to be deployed under supervised conditions by trained personnel. Here, we develop and validate the Comprehensive Online Sleep Monitoring Scale (COSMOS), a self-reported sleep scale optimised for online, independent administration. Using data from N = 5,815 adults, we show that COSMOS has good internal (Cronbach’s α = 0.85) and convergent validity, via strong associations with established sleep scales. Poorer COSMOS sleep scores were found in participants diagnosed with psychological conditions, more frequent depression or anxiety symptoms, and higher compulsivity or neuroticism traits, demonstrating good construct validity. We propose COSMOS as a comprehensive and validated sleep assessment that is suitable for large-scale online transdiagnostic and mental health research.

## Introduction

Sleep is a fundamental physiological process that plays an integral role in maintaining health and wellbeing (Foster, 2020; Ramar et al., 2021; Watson et al., 2015). Contrary to the conventional view of sleep problems being secondary symptoms of other conditions, emerging studies highlight that they can be associated with subsequent neuropsychiatric and neurological diagnoses (Freeman et al., 2020). For instance, Rapid Eye Movement Behaviour Disorder (RBD) is strongly associated with later emergence of neurodegenerative disorders including Parkinson’s disease and Dementia with Lewy Bodies (increased risk of around 90%; Galbiati et al., 2019). In psychosis, major depressive disorder, and anxiety, sleep disturbances are commonplace and may, in some cases, also predate the onset of new episodes (Ferrarelli, 2020; Waite et al., 2020). The relevance of sleep extends beyond clinical conditions to the general population, with sleep consistency and morning preference being linked to better academic performance (Hershner, 2020), whereas shorter sleep durations and elevated disturbances are associated with risk of injury at work and lower productivity. Sleep disruption also greatly impacts memory consolidation and learning processes (Ashton & Cairney, 2021; Gutiérrez et al., 2024; Guttesen et al., 2023). Consequently, the examination of sleep has become widespread in psychological, psychiatric and neurological studies (Kucharczyk et al., 2012).

Sleep is complex, multistage and can be disrupted in a variety of ways. Studying sleep often necessitates the use of multiple measurement instruments designed to capture different relevant constructs. Critically, some of the most common scales were originally designed for deployment under supervised conditions by trained personnel or those with clinical expertise. This presents two key limitations to their utility. First, standard scales may not be ideal for deployment by non-sleep specialists engaged in research (Klingman et al., 2017). More importantly, there is a growing application of assessment scales online via peoples’ home devices – e.g., personal computers, tablets, and smartphones. This shift to online deployment has been motivated by the scalability, repeatability, accessibility and affordability relative to supervised assessment methods, presenting an opportunity to study variability in neurological, psychiatric and mental health conditions in previously unachievable population size and detail (Bălăeţ et al., 2024; Binoy et al., 2024; Hampshire et al., 2020). However, a barrier to realising this potential is that scales designed for supervised deployment can produce different score distributions when deployed online (Morton et al., 2020); therefore, they require either online validation and recalibration or specific development taking into consideration the online medium they are to be deployed in.

The ubiquity of digital technologies also has direct relevance to sleep problems; specifically, the dynamics of sleep-related behaviours have been evolving in response to the increasing centrality of technology in our daily lives. Extensive use of personal and home technological devices such as mobile phones, tablets, televisions, and e-readers can substantially affect sleep behaviours and the quality of sleep one experiences (Liu et al., 2019; Scott & Woods, 2019). Therefore, current sleep assessments must adapt not only for deployment through these technologies but also to measure their influences on sleep.

Here, we developed a novel and comprehensive questionnaire designed to be suitable for modern online assessment of sleep. The questionnaire targets constructs from popular sleep scales and extends beyond those constructs with unique items that specifically address sleep-related behaviours resulting from the accessibility of technological devices. Crucially, we specifically tailored the wording of items to be unambiguous when deployed under unsupervised conditions, then validated them through deployment on an internet– and app-based research platform (Cognitron).

We intended that this Comprehensive Online Sleep Monitoring Scale (COSMOS), would measure eight key facets of sleep. These included (1) sleep disturbance, (2) daytime dysfunction, (3) symptoms indicative of sleep disorders, (4) bedtime routines, which included extent of screen time before bed, (5) medication use to support sleep, (6) chronotype, or natural preference to be active in the morning or evening, (7) sleep latency and duration, and (8) sleep quality. We expected COSMOS to show good, convergent validity when compared to established and widely used scales including the Pittsburgh Sleep Quality Index (PSQI; Buysse et al., 1989), Morningness-Eveningness Questionnaire (MEQ; Horne & Ostberg, 1976), and Epworth Sleepiness Scale (ESS; Johns, 1991).

Given the intended application of this sleep scale in future psychiatry research, and to investigate the scale’s construct validity, we explored the relationship between COSMOS and psychiatric conditions, psychological symptoms and risk-associated traits previously linked with sleep disruptions (Freeman et al., 2020). We hypothesised that worse sleep scores, as measured by COSMOS, would be evident in people reporting one or more pre-existing mental health diagnoses and those with elevated frequencies of depression or anxiety symptoms as measured using items from the nine-item Patient Health Questionnaire (PHQ-9; Kroenke et al., 2001) and Generalized Anxiety Disorder-7 (GAD-7; Spitzer et al., 2006), since sleep disturbances are common symptoms of both conditions. Compulsivity is characteristic of several psychiatric disorders, particularly obsessive-compulsive-related disorders, in which sleep disturbances are prevalent (Segalàs et al., 2021; Tiego et al., 2023). We expect that higher compulsivity as measured by the Cambridge–Chicago Compulsivity Trait Scale (CHI-T; Chamberlain & Grant, 2018; Tiego et al., 2023) will be associated with higher scores on COSMOS. We also sought to assess the relationship of COSMOS measures with neuroticism as measured by an abbreviated version of the Big Five Personality Inventory (Gosling et al., 2003; John, 1991), as this personality trait has been proposed as a potential risk factor across psychiatric conditions (Kotov et al., 2010; Lahey, 2009; Ormel et al., 2013; Schirmbeck et al., 2015) and has been identified as an independent risk factor for sleep problems (Akram et al., 2023). We benchmarked the discriminative sensitivity of COSMOS relative to established sleep scales when assessing their associations with the above conditions, symptoms, and traits.

## Methods

### Participants, Design, and Procedure

Twenty-eight thousand seven-hundred fifty-two (28,752) participants who had taken part in a follow-up cognitive assessment at the end of 2022 as part of the Great British Intelligence Test (GBIT; Hampshire, 2020) on the Cognitron testing platform were invited to this study. Five-thousand eight-hundred fifteen complete responses were recorded. For full demographics see **Table 1**.

**Table 1.**
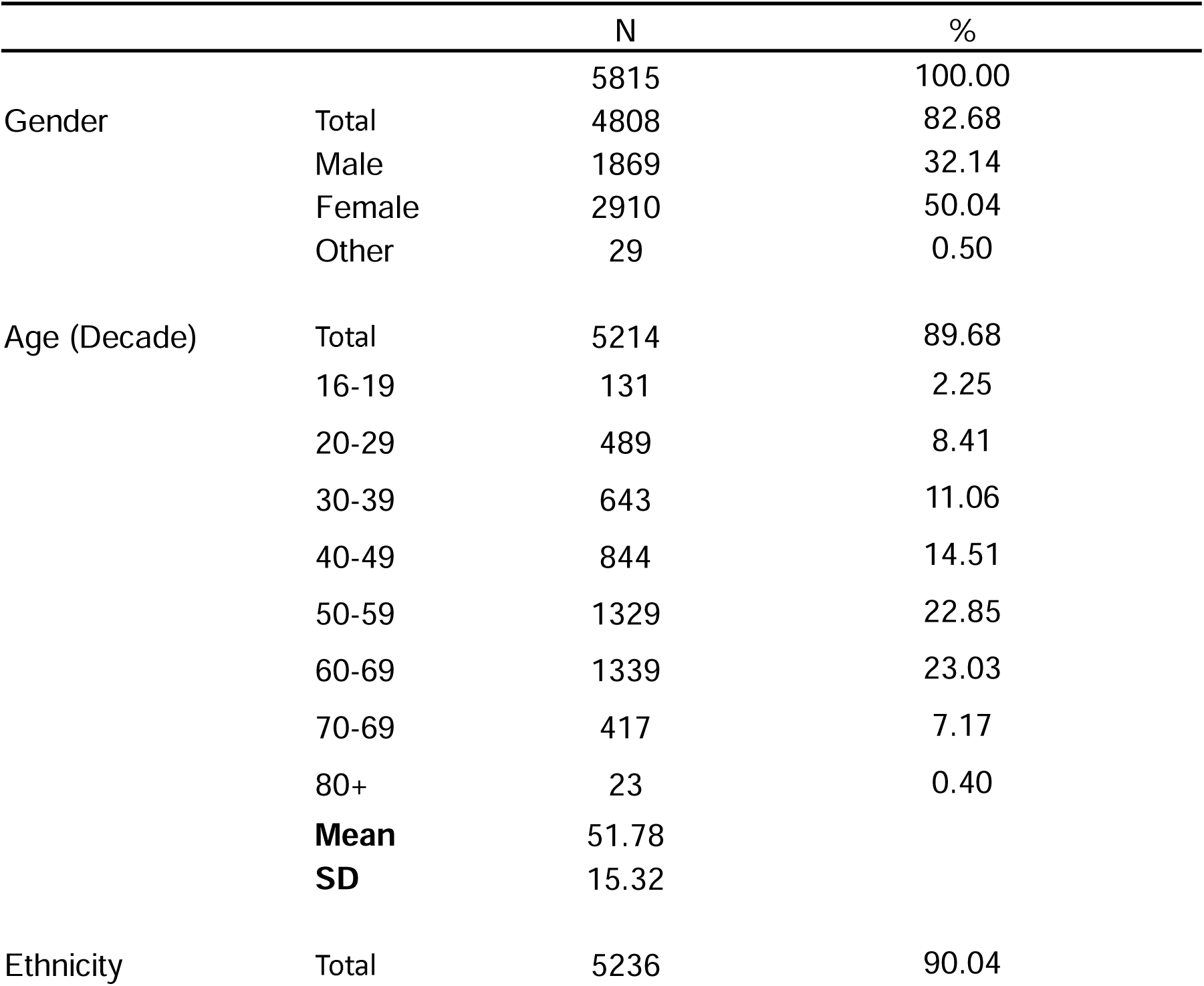
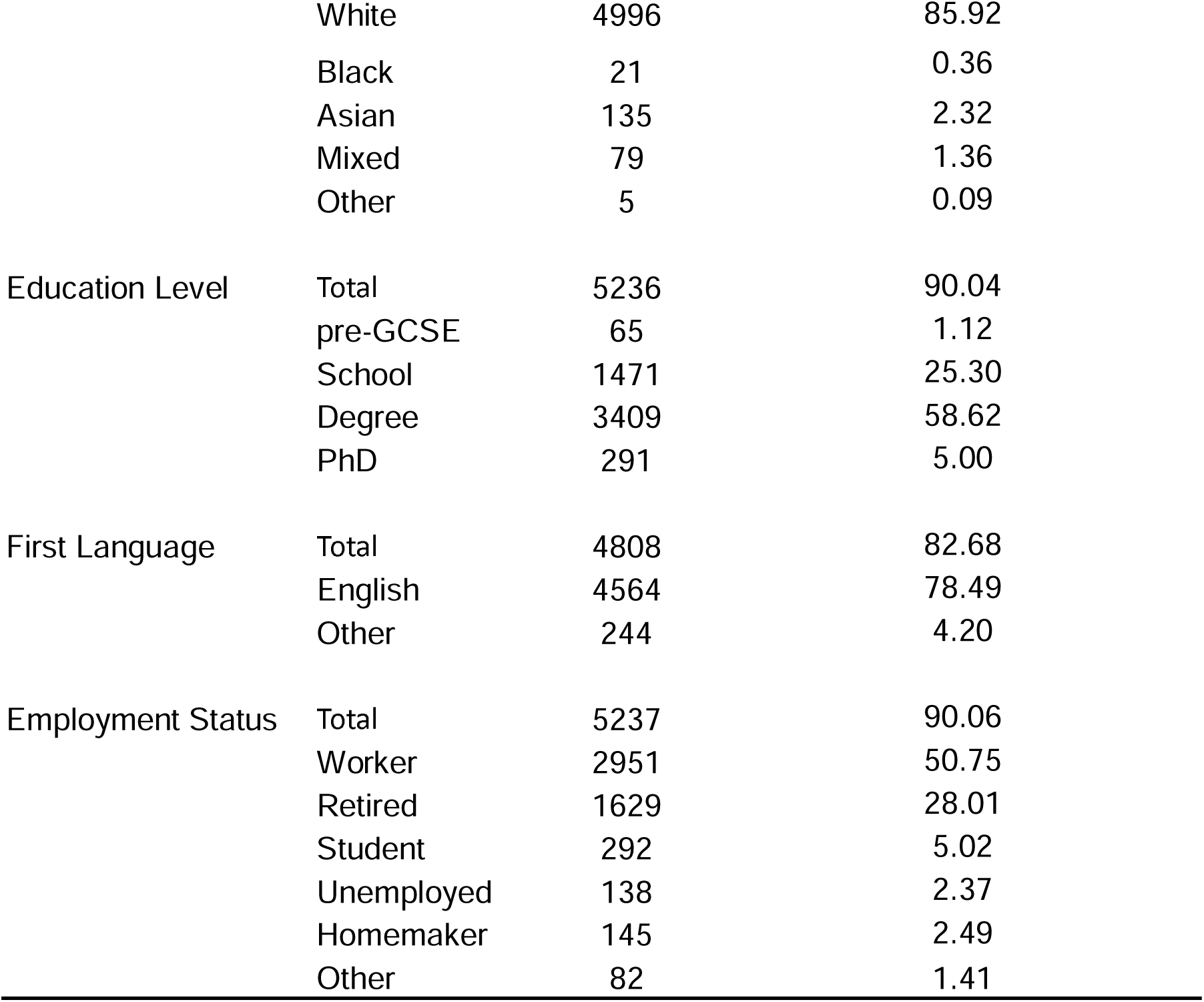
Demographics.

COSMOS, PSQI, ESS, and MEQ were presented to all participants in this fixed order, with visible scale titles. Participants could complete the questionnaire on their personal devices (e.g., mobile phone, tablet, laptop, personal computer) and browser of choice.

## Materials

### Sleep Metrics

#### COSMOS Development

The most common sleep assessment scales were compiled based on a literature search in PubMed using the following terms: (sleep quality[Title]) AND ((psychometric*[Title]) OR (questionnaire[Title]) OR (scale[Title]) OR (index[Title]) OR (diagnos*[Title]) OR (instrument[Title])). After filtering for studies conducted with human samples, the number of search results was narrowed down to 194 from 241.

The papers’ relevance was evaluated based on their titles and the scales used or mentioned. Of the eighteen unique scales that were identified, thirteen were deemed relevant based on a deeper inspection of the literature. Scales that were sample-or disorder-specific were removed.

Items from the thirteen scales carried forward were evaluated to identify common themes and items (see **Appendix A** for full list of scales). Items common across multiple scales were collapsed and the frequency of the item was noted. The items were then grouped based on these broad themes and further categorized based on the specific subject of the item. For example, item variants on the subject “Do you snore (loudly)?” were found in seven of the scales and classified as “breathing issues” and then subcategorized under “snoring”. This narrowed down the questions from 334 to 74 items.

A shortlist of relevant subjects and questions to include was created based on the number of items in each category, how common items were across the scales, and the relevance of the items to assessments of sleep in the general population. This yielded 49 items of which 10 were removed due to having too much overlap in content. A list of 31 essential items was created and reworded to fit with the intention of unsupervised online administration and response within the web browser.

After consultation with researchers and sleep experts, items were added, removed, and modified. A “bedtime habits” subscale was added to capture common behaviours around sleep primarily covering technology use, reading, and prayer or meditation. More detail around drug or medication use to support sleep was added to encompass illegal drugs, cigarette smoking, and alcohol consumption as well as prescribed and over-the-counter medications. These were phrased in the context of use to aid sleep rather than general consumption. Questions about the quantity of use were added. Additional sleep onset and wake-up items were added to capture any disparity in sleep duration and onset from weekdays to weekends, also known as social jetlag. Finally, three free text, or open-ended, items were added to allow participants to share, in their own words, details about any medical conditions that may affect sleep quality, sleep problems experienced in the past four weeks, and if their sleep over the past four weeks differed from the past few years and why (see **Figure 1** for a summary of the scale and study design).

**Figure 1.**
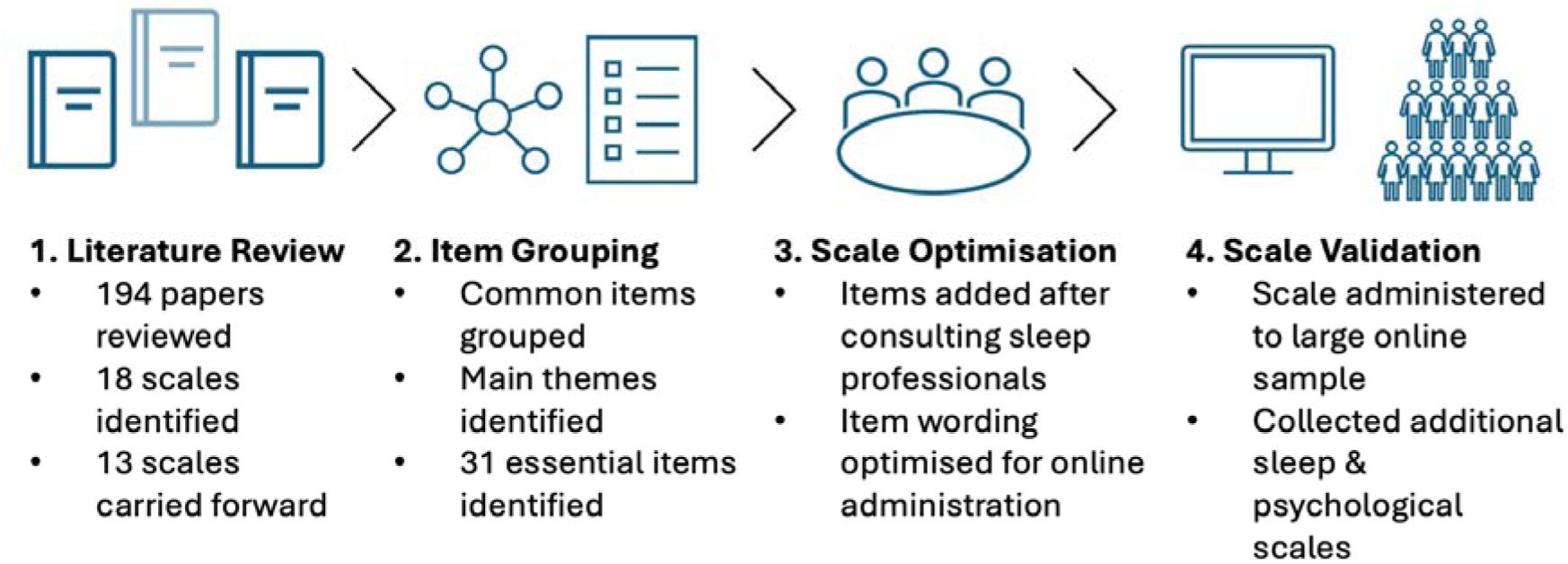
Scale Development Process.

The final scale (**Table 2**) comprised a total of 49 compulsory questions that were displayed to every participant and 17 nested items that were only completed conditional on the response to the previous question. For instance, participants who indicated they never took naps did not receive questions about the frequency, duration, and spontaneity of their naps.

**Table 2.**
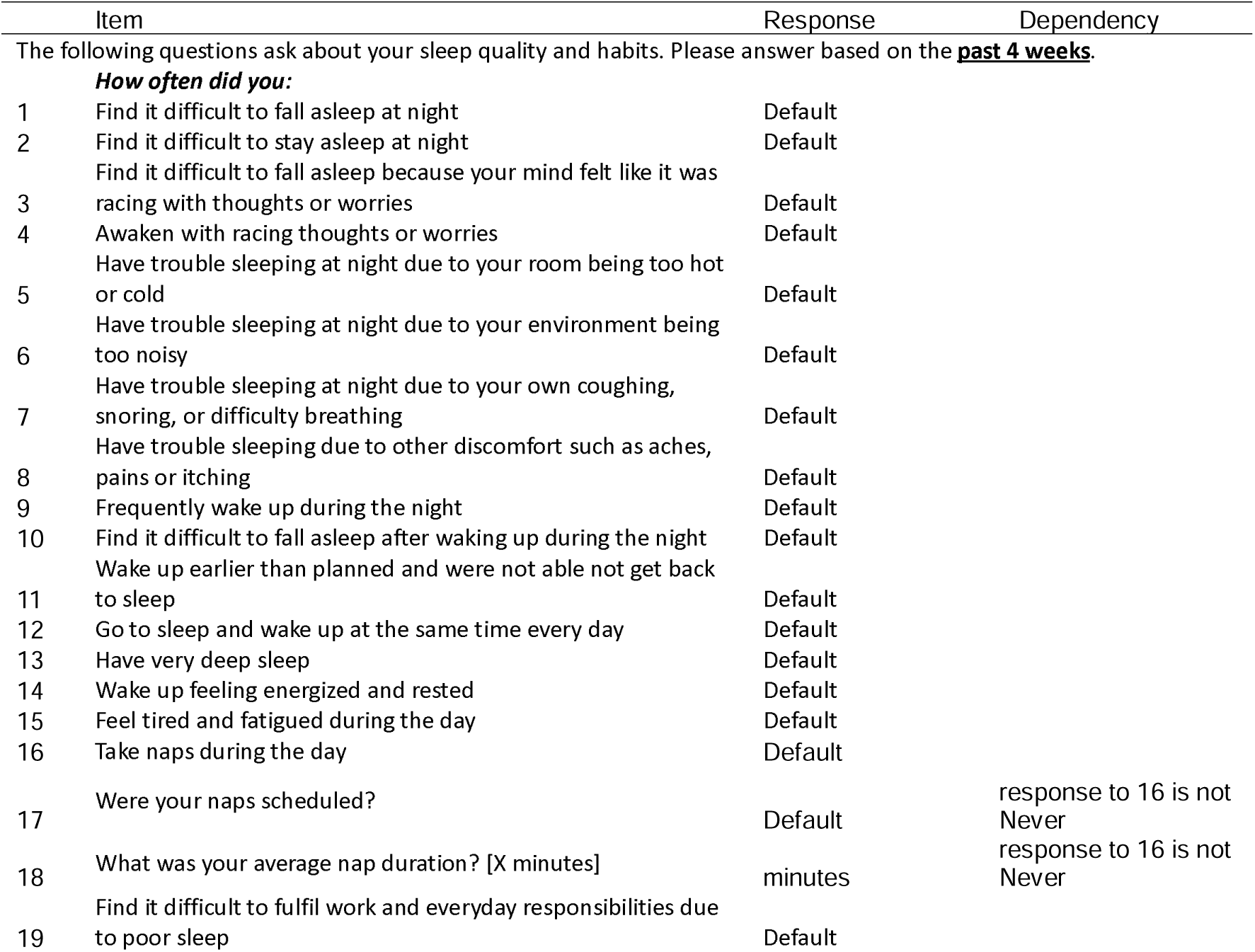
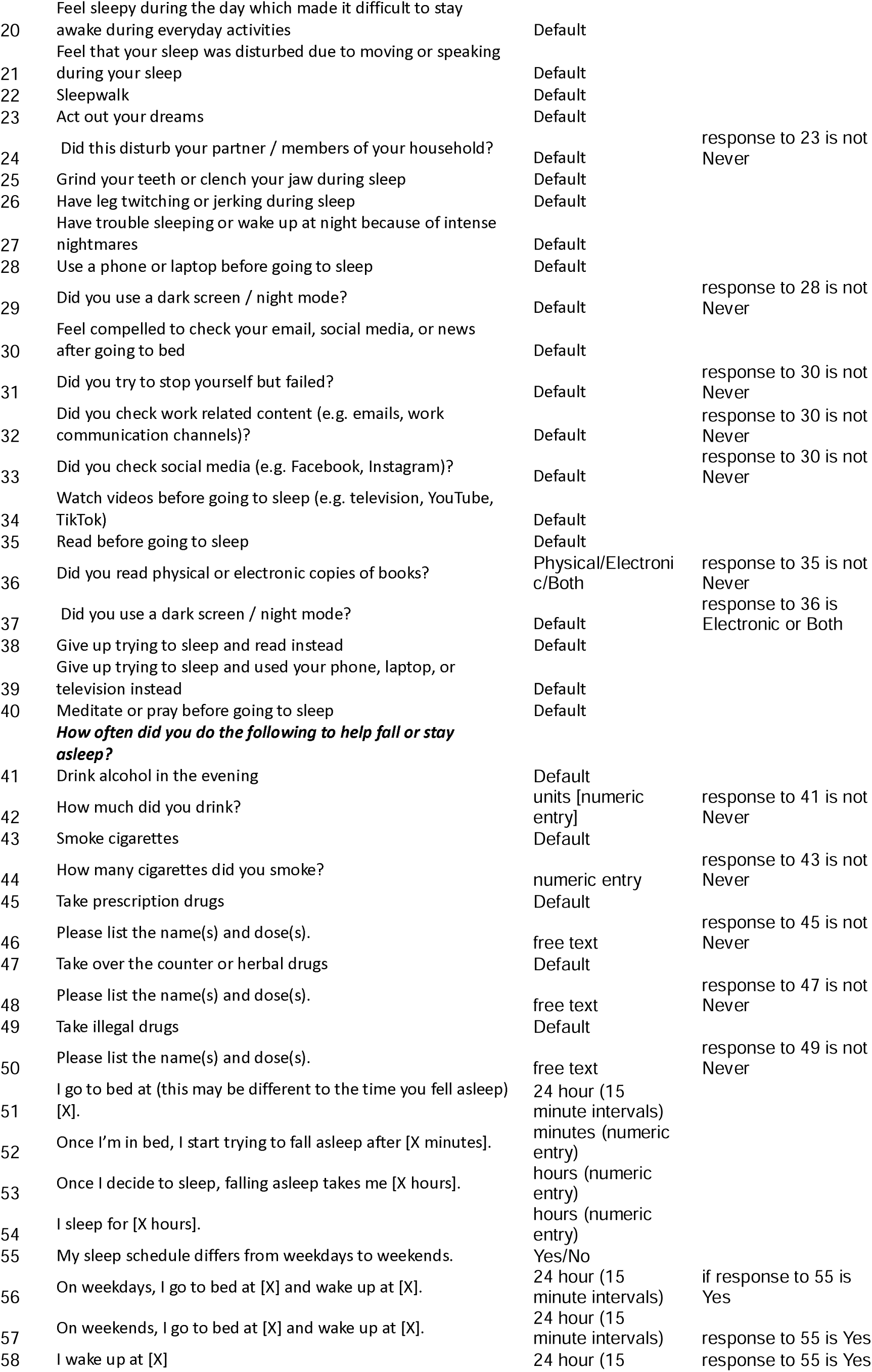
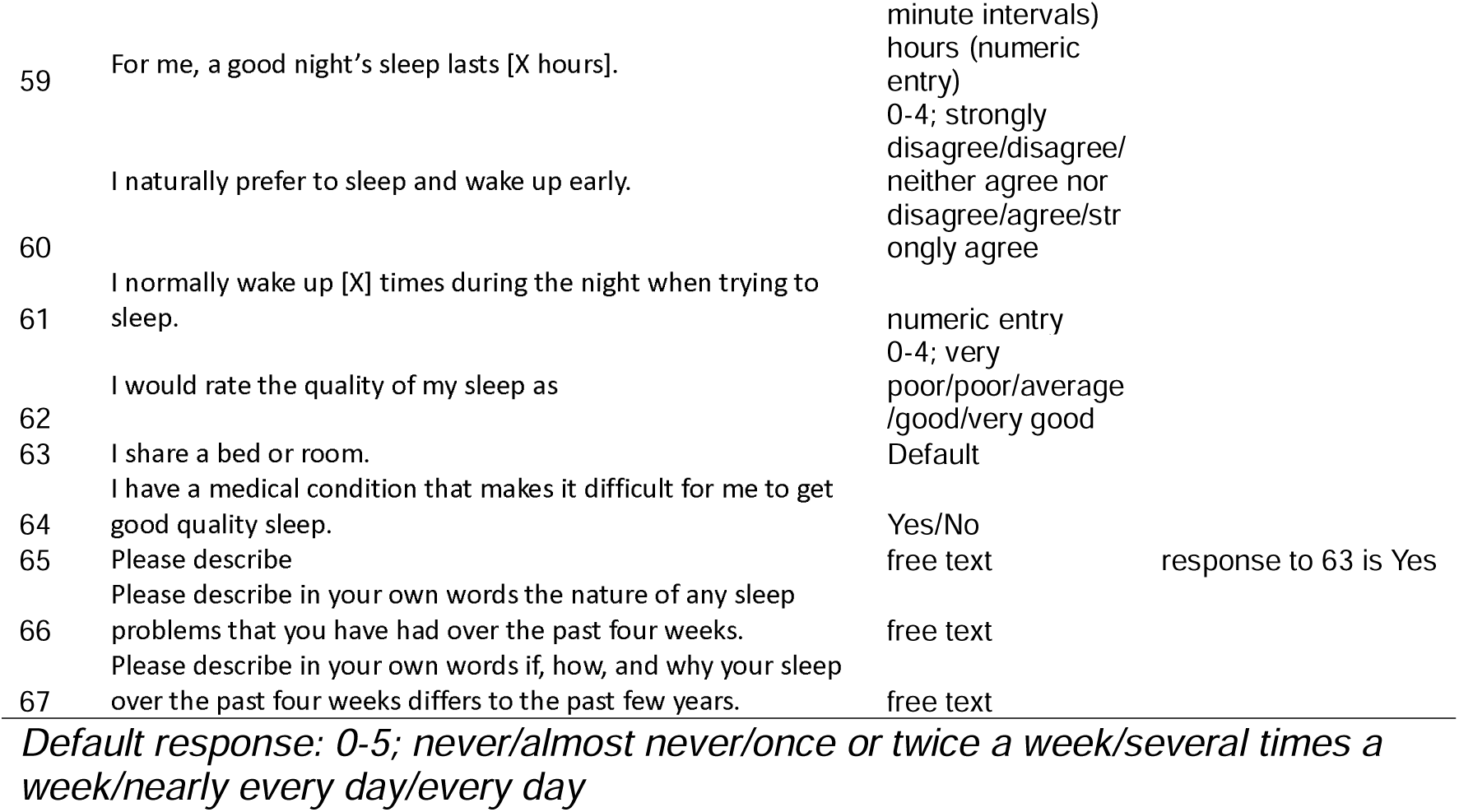
COSMOS Items.

The questionnaire specified that responses should be based on the past four weeks. The majority of items were scored on a six-point Likert scale from zero, or “never”, to five, “every day”. Exceptions to this included some nested items, where a more specific response was appropriate (e.g. yes / no questions, quantities and dosage, and screen time specifications), timings such as sleep onset or duration, free text items, a sleep quality item that asked for a rating of sleep quality on a five-point Likert scale ranging from “very poor” to “very good”, and a propensity to morning chronotype item which was rated on a five-point Likert scale ranging from “strongly disagree” to “strongly agree”.

#### PSQI

The PSQI (Buysse et al., 1989) was used to obtain a measure of sleep quality and its components more generally. The scale generates a global sleep quality metric alongside scores for seven components of sleep quality including, self-rated sleep quality, sleep latency, sleep duration, sleep efficiency, sleep disturbances, daytime dysfunction, and medication use around sleep. The scores for each component range from zero to three and these scores are then added, resulting in a single global score from 0-21 with higher scores indicating poorer sleep quality. The scale has been used widely for research and has good reliability according to the reported Cronbach’s (1951) alpha (Cl) of 0.83.

#### MEQ

Chronotype, or circadian rhythm preference, was measured using the MEQ (Horne & Ostberg, 1976). Here the respondent is asked to indicate the times they feel most alert or tired as well as their sleep-wake preferences. Responses were on a five-point Likert scale. Scores were calculated based on the total score for each item after reverse coding respective items such that higher scores indicated morning-type groups and lower scores evening-type individuals. The scale is widely used and reliable; Cronbach’s Cl of 0.87.

#### ESS

To assess daytime dysfunction – or sleepiness – we utilized the ESS (Johns, 1991). The ESS measures subjective sleepiness, Cronbach’s Cl of 0.88. The scale presents eight everyday situations where the respondent is asked to rate their likelihood to doze off on a four-point Likert scale from “no chance of dozing” to “high chance of dozing.” Responses are added to produce a total score from 0 to 24, with a score of 24 indicating excessive sleepiness.

### Mental Health Diagnoses

The presence of a neurological or psychiatric illness was assessed through self-report. If respondents indicated they had been diagnosed with a neurological condition, psychiatric condition, or both, they were provided with a list of relevant diagnostic labels to select from. For psychiatric disorders, respondents could choose from Depression, Anxiety, Attention Deficit Hyperactivity Disorder, Obsessive Compulsive Disorder, Bipolar Disorder, or other. Multiple options could be selected at once. Respondents that indicated they did not have any neurological or psychiatric diagnoses were not presented with these two follow-up questions.

### Depression and Anxiety Symptoms

Depressive and anxious symptoms were measured using a subset of items from the PHQ-9 (Kroenke et al., 2001) and full GAD-7 (Spitzer et al., 2006). To ensure suitability for the general population, specific items from the PHQ-9 related to appetite, self-worth, agitation, and suicidality were excluded.

### Compulsivity and Big Five Personality traits

The CHI-T (Chamberlain & Grant, 2018), a short comprehensive compulsivity metric, was utilised to capture compulsive traits. The scale has two latent factors, one relating more to perfectionism and the second to compulsive soothing behaviours (Tiego et al., 2023). Some studies have also reported a three factor model, with perfectionism, cognitive rigidity, and reward drive, where cognitive rigidity and reward drive are included here under compulsive soothing (Hampshire et al., 2021).

An abbreviated version of the Big Five Inventory (John, 1991) was administered to capture personality traits, primarily neuroticism. The scale has five established factors – openness, agreeableness, extraversion, conscientiousness, and neuroticism – where the combination of scores on each describes an individual’s personality traits.

## Analysis

### Tools and Software

Confirmatory factor analyses were conducted in R version 4.2 (R Core Team, 2022) using the Lavaan package (Rosseel, 2012). All other data processing and analyses were conducted using Pandas in Python version 3.9 (The Pandas Development Team, 2022).

Pingouin (Vallat, 2018) was used to run ANOVAs and respective post hoc tests, Statsmodels (Seabold & Perktold, 2010) to test multiple regression models, Scipy (Virtanen et al., 2020) to generate correlations and associated *p*-values, and FactorAnalyzer (Biggs, 2022) to create fitted exploratory factor models from which individual factor scores were extracted.

### Pre-Processing

All items were converted from string responses to Boolean or scaled responses. Items were appropriately reversed such that all higher scores were indicative of poorer sleep behaviour or quality. Any missing data or skipped questions were coded as “not a number” (“NaN”). Free text items were dropped from further analyses described here. No imputation of missing variables was applied in the analyses. Scores were all z-scored.

### Scoring

#### COSMOS

Social jetlag was calculated by taking the difference in sleep duration on weekends from weekdays. These sleep durations were calculated using the weekday and weekend bedtime and wake-up time items (items 56-59, **Table 2**). Those who indicated no difference in their sleep during the week and weekend received a score of zero. These items were then dropped from further analyses for all participants.

#### Depression and Anxiety Symptom Scales

A total score was calculated for depressive symptoms by taking the sum of the subset of PHQ items administered. The same was done using the full GAD-7 to calculate a total score for symptoms of anxiety.

#### Compulsivity and Neuroticism Scales

To calculate scores for the CHI-T compulsivity factor, exploratory factor analysis (EFA) was used to extract factor scores for the two CHI-T factors, as per Tiego et al. (2023). Factors were extracted using principal axis factoring and a varimax rotation was applied. Individual scores for each factor were then calculated using the fitted factor model. The compulsive soothing factor was taken forward for analysis rather than perfectionism as the items under the compulsive soothing but not perfectionism factor has been associated with poorer functional outcomes (Hampshire et al., 2021; see **Appendix B** for full EFA results).

Principal axis factoring set to five factors with varimax rotation was applied to the Big Five Inventory to extract factor scores. Neuroticism scores were multiplied by –1 such that higher scores indicated higher levels of neuroticism and then were carried forward for analysis (see **Appendix C** for full EFA results).

## Results

### Internal Structure

The full item COSMOS showed good internal consistency as indicated by a Cronbach’s Cl of 0.85. A confirmatory factor analysis (CFA) was conducted to test the hypothesized eight-factor structure of COSMOS. Nested variables were defined in the model as additional covariances. The model showed reasonable fit, χ*^2^*(1666) = 31026.86, *p* < .001, CFI = .81, RMSEA = .06. See **Appendix D** for full factor loadings.

### Convergent Validity

A multistep approach was taken to assess the extent to which COSMOS aligned with established sleep questionnaires. First, a second CFA of COSMOS was conducted using the hypothesized factor structure but separating the sleep latency and duration factor into four distinct factors: sleep latency, sleep duration, sleep onset, and social jetlag. This was done to make the COSMOS factor structure more similar to the structure used in the established scales. The model showed a slightly poorer fit to the original eight-factor model, χ*^2^*(1368) = 25089.80, *p* < .001, CFI = .78, RMSEA = .06. See **Appendix E** for full factor loadings.

Next, a CFA of the established scales was conducted using a nine-factor model. These were made up of the seven predefined PSQI factors, daytime dysfunction which included the ESS items, and a chronotype factor containing the MEQ items. The model showed a similar fit quality to COSMOS models, *^2^*(399) = 9274.80, *p* < .001, CFI = .80, RMSEA = .06. See **Appendix F** for full factor loadings.

These fitted models of COSMOS and established scales were used to extract individual latent variable scores for each of the factors. A Pearson correlation with a Bonferroni correction for multiple comparisons was calculated for each pair of scores to assess the relationship between the subscales (**Figure 2**). This showed statistically significant correlations of moderate to strong effect size between the COSMOS subscales and the corresponding established subscales. The correlation between medication subscales was statistically significant but weak.

**Figure 2.**
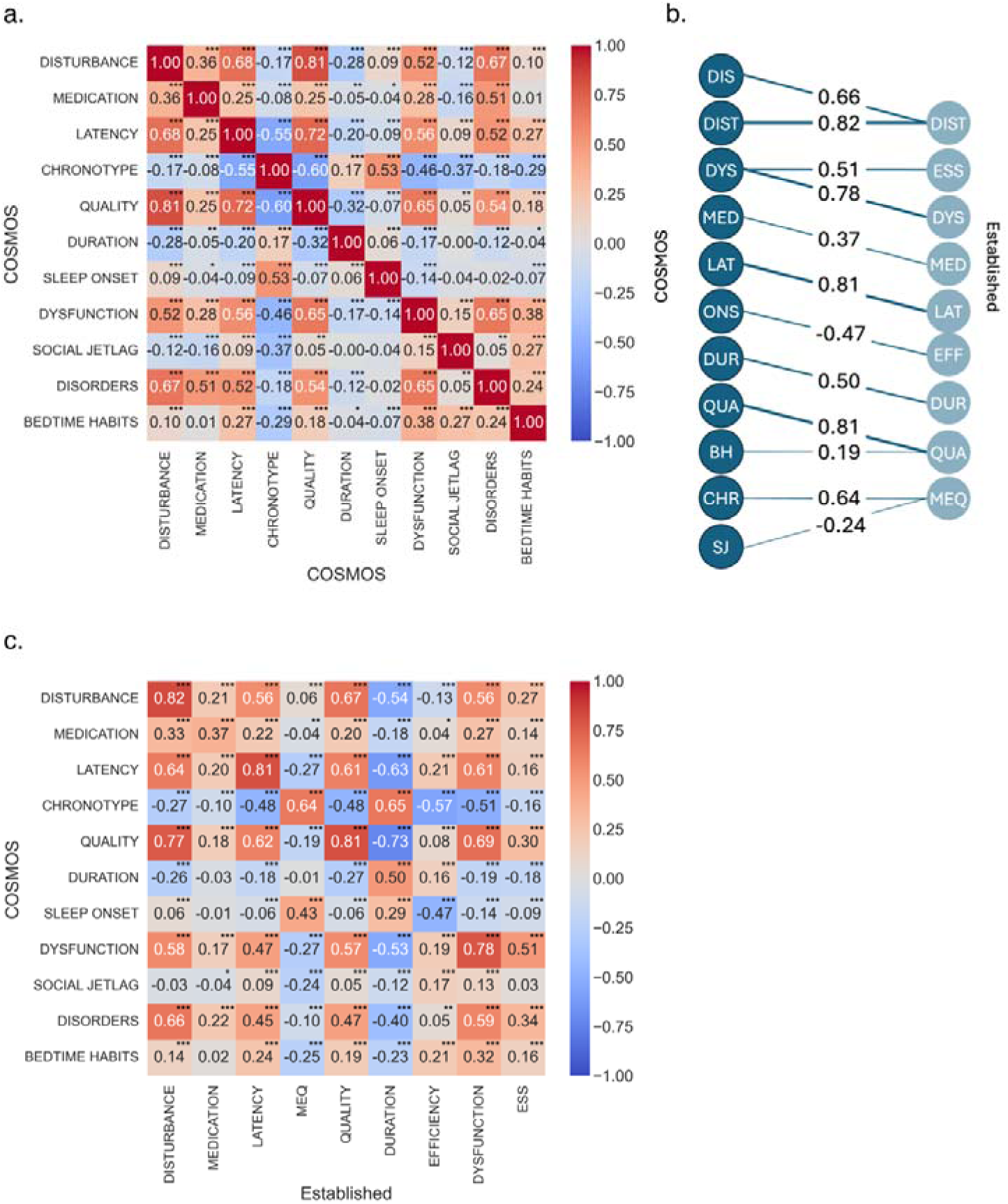
COSMOS Latent Variable Score Correlations. a. COSMOS latent variable model subscale score correlations and b. corresponding variable correlations between COSMOS (left) and the established scale latent variable models (right) c. an asymmetric correlation matrix describing the correlations between corresponding scales across COSMOS and the established model. p-values all corrected for multiple comparisons using Bonferroni correction, \*\*\**p* < .001, \*\**p* < .01, \**p* < .05.

The social jetlag, disorders, and bedtime habit subscales did not have clear corresponding established subscales to be compared to in this study. Social jetlag scores did not correlate well with any of the subscales but showed a significantly weak negative correlation to the MEQ. Similarly, the bedtime habits subscale was weakly but significantly correlated with timing-related subscales including the MEQ, latency, and efficiency and the dysfunction subscale.

## Associations

### Self-Report Psychiatric Diagnostic Labels

A mixed ANOVA was conducted to assess sleep differences across the COSMOS subscales between participants who had no psychiatric conditions (n = 5024) and those who reported having at least one psychiatric, but no neurological conditions (n = 506).

Participants who indicated having a neurological condition were excluded from this analysis (n = 285). See **Table 3 for included disorders.**

**Table 3.** Number of participants in each diagnostic group.

| Diagnosis | Count |
| --- | --- |
| None | 5024 |
| Depression | 330 |
| Anxiety | 89 |
| Obsessive Compulsive Disorder | 36 |
| Other | 32 |
| Attention Deficit Hyperactivity Disorder | 19 |

There was a significant main effect of self-reported psychiatric diagnosis on total COSMOS scores, *F* (1, 5528) = 250.46, *p* < .001, η_p_^2^ = .043. Pairwise post-hoc tests were conducted with a Bonferroni correction of multiple comparisons. These showed significantly higher scores on all COSMOS subscales, except for the Chronotype subscale where this effect was reversed (**Table 4**). The mean differences were significant for every subscale except duration (**Figure 3**). The effect sizes ranged from small to large, with the dysfunction subscale showing the largest effect.

**Figure 3.**
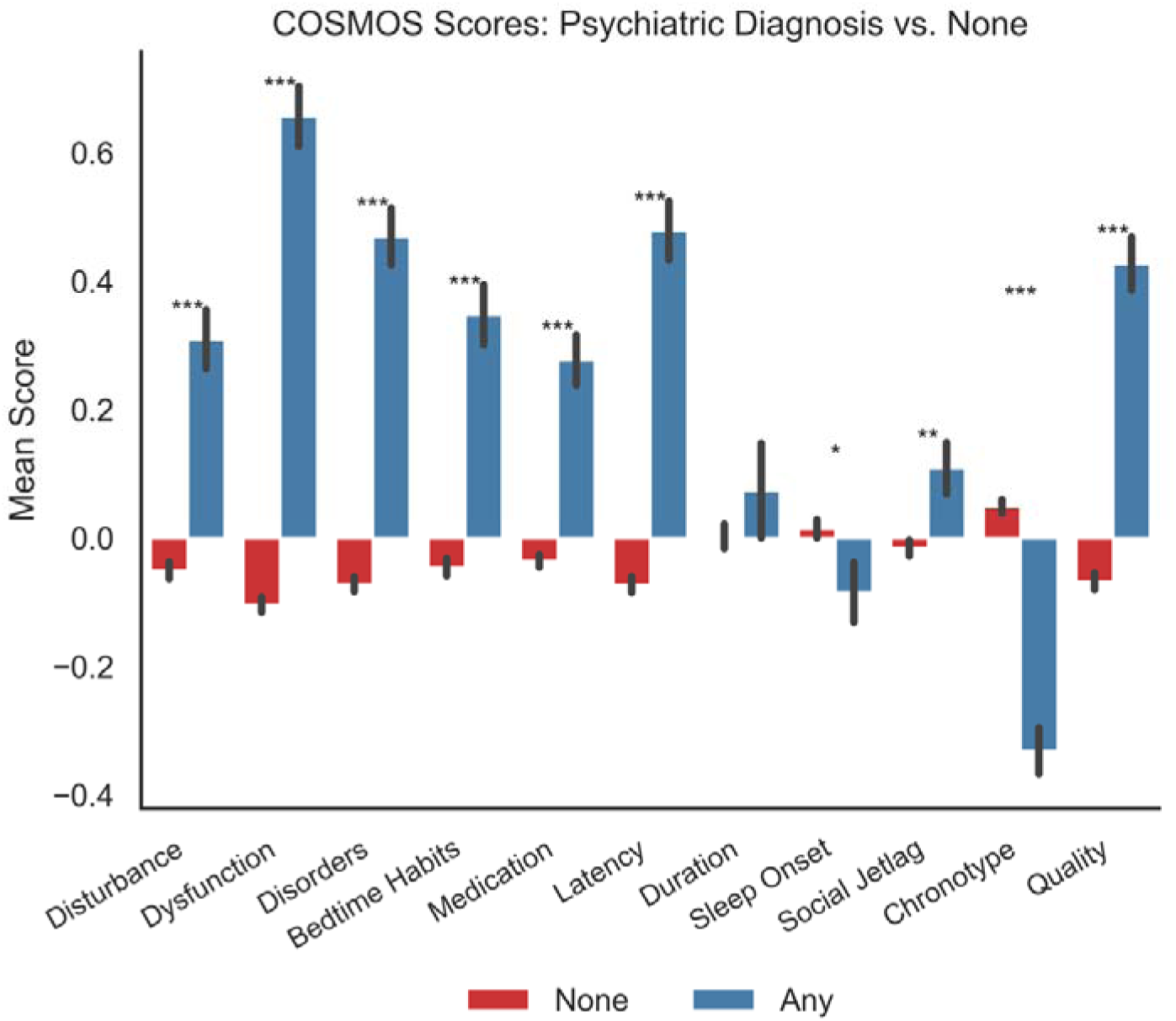
Difference in COSMOS subscale latent variable scores between participants diagnosed with any or no psychiatric condition. Post-hoc test of z-scored latent variable scores for each COSMOS subscale with Bonferroni correction for multiple comparisons, \*\*\**p* < .001, \*\**p* < .01 \**p* < .05.

**Table 4.** Mixed ANOVA post-hoc results subscales x psych group with *FDR corrected p-values and Cohen’s d*.

| Subscale | *t* (df) | *p* | *d* |
| --- | --- | --- | --- |
| Disturbance | -7.55 (590.8) | <.001 | -0.38 |
| Dysfunction | -15.66 (572.34) | <.001 | -0.87 |
| Disorders | -11.89 (572.47) | <.001 | -0.66 |
| Bedtime Habits | -8.12 (589.18) | <.001 | -0.42 |
| Medication | -7.75 (574.43) | <.001 | -0.43 |
| Latency | -11.5 (580.91) | <.001 | -0.61 |
| Duration | -0.94 (574.08) | .349 | -0.05 |
| Sleep Onset | 2.04 (600.95) | .046 | 0.10 |
| Social Jetlag | -2.98 (610.45) | .004 | -0.14 |
| Chronotype | 10.1 (598.11) | <.001 | 0.50 |
| Quality | -11.35 (605.04) | <.001 | -0.54 |

### Depression and Anxiety Symptoms

The relationships between COSMOS and established anxiety and depression symptom scales were assessed using multiple linear regressions. Separate multiple linear regression models were run with COSMOS latent variable scores as the independent variables and either the depression or anxiety scores as the dependent variable. The same analyses were conducted using the latent variables extracted from the established scales’ CFA as the independent variables. Full results can be found in **Appendix G**.

The COSMOS and established scale models were both statistically significant and explained over half of the variance in depression scores (*F*(11, 5225) = 556.80, *p* < .001, *R*^2^ = 0.54; *F*(9, 5227) = 661.70, *p* < .001, *R*^2^ = 0.53, respectively). The COSMOS dysfunction subscale showed a large effect size followed by small effect sizes for the bedtime habits and latency subscales (**Figure 4**).

**Figure 4.**
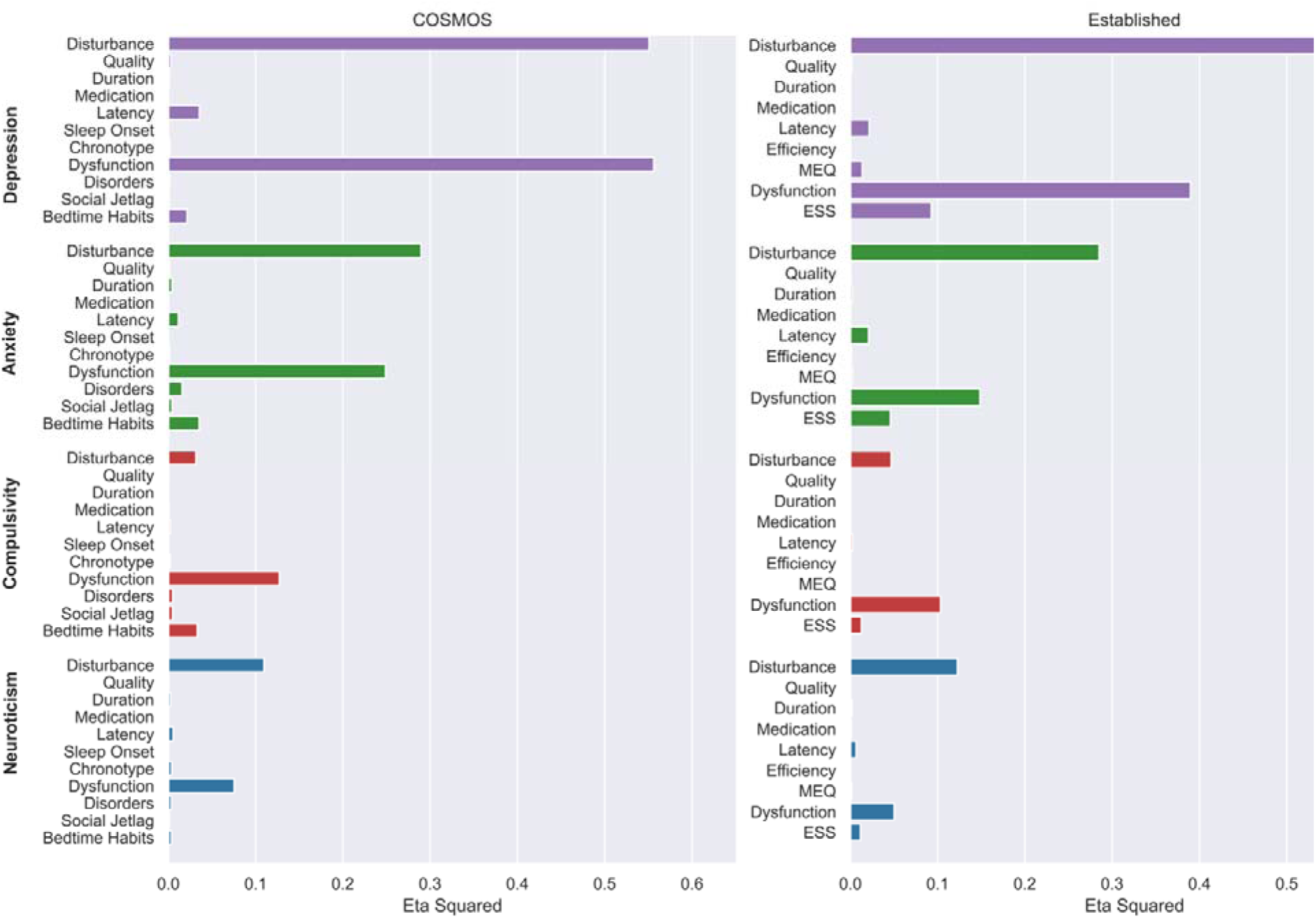
Effect sizes of regression coefficients for COSMOS and established scale subscales when estimating psychological symptoms and traits.

COSMOS and established scales both significantly explained over 30% of the variance in anxiety scores, with COSMOS explaining 38% (*F*(9, 5227) = 290.80, *p* < .001, *R*^2^ = 0.38) and established scales 34% (*F*(9, 5227) = 293.60, *p* < .001, *R*^2^ = 0.34) of the variance. Again, the dysfunction subscale contributed most to increases in GAD scores as shown by its large effect size along with disorders, bedtime habits and latency which had small effect sizes (**Figure 4**).

### Personality Traits

Separate multiple linear regression analyses were conducted between the COSMOS subscales and extracted CHI-T compulsivity and BIG-5 neuroticism factors. Identical analyses were conducted with the established scales’ factor scores as the independent variable. COSMOS significantly predicted scores on the compulsive soothing factor, explaining 18% of the variance (*F*(11, 5225) = 98.31, *p* <.001, *R*^2^ = 0.18). This was similar for the established scales, (*F*(9, 5227) = 95.97, *p* <.001, *R*^2^ = 0.14). COSMOS’s dysfunction subscale had the greatest contribution, with a medium effect size (**Figure 4**). The bedtime habit and social jetlag subscales’ contributions had small effect sizes (**Figure 4**).

The model using COSMOS subscales to predict scores on the extracted neuroticism factor was statistically significant ( *F*(11, 5224) = 97.28, *p* <.001, *R*^2^ = 0.17) and this was very similar to that of the established scales (*F*(9, 5226) = 113.10, *p* <.001, *R*^2^ = 0.16). The dysfunction subscale, again, contributed most to this, with a medium effect size (**Figure 4**). See **Appendix G** for the full set of results. Repeating the analyses but with independent models for each predictor produced the same pattern of results but with larger effect sizes (**Appendix H**), likely due to feature collinearity.

## Discussion

There is great potential utility in being able to assess sleep quality and its components comprehensively and remotely without supervision, including deployment longitudinally at a large population scale. COSMOS was developed to address these needs and, here, we demonstrate that it has good convergent validity with popular established scales and equivalent or stronger associations to psychiatric conditions, mental health symptoms and personality traits.

When applying a confirmatory approach to examine the internal structure of COSMOS, where items were assigned to factors according to the extant sleep literature, and taking inspiration from the established scales used in this study, the model provided a comparable fit for the data as those established scales. Furthermore, COSMOS showed high internal consistency as measured by Cronbach’s alpha. Corresponding scores from COSMOS and established scales correlated well. Taken together, these results demonstrate good validity of the constructs COSMOS was intended to measure.

COSMOS was created to be a comprehensive tool for current times; therefore, it includes some elements not captured by the established PSQI, ESS, or MEQ scales. For example, medication in the PSQI includes over-the-counter or prescribed medication to support sleep whereas COSMOS does so for *any* drug or substance use in the context of aiding sleep, including alcohol, cigarettes, and illicit substances, over and beyond just over-the-counter and prescription medications. This added variance could explain the weaker correlation observed between these two subscales, particularly as some of the additional substances, such as alcohol, could impair sleep despite the intended use as a sleep aid (He et al., 2019). Another consideration is that this scale was administered in the United Kingdom where cannabis use remains illegal. Therefore, this information may be unintentionally categorised under “over-the-counter” or “prescription medication” if the scale was administered in a country where cannabis use was legal. For a more generalisable global scale, future iterations may benefit from explicitly addressing cannabis or listing it as an independent item.

The instrument showed good construct validity, with poorer COSMOS sleep scores in participants diagnosed with psychological conditions, more frequent depression or anxiety symptoms, and higher compulsivity or neuroticism traits. We evaluated the associations between COSMOS and specific aspects of mental disorders including the presence of any disorder as well as scores relating to depression and anxiety. Links between COSMOS and compulsivity, as well as Big Five personality traits were evaluated. The relevance of sleep in understanding psychiatric disorders has been well-established, a significance that grows as we move towards a transdiagnostic approach in psychiatric diagnosis and treatment (Freeman et al., 2020). Given this emerging perspective, we sought to create a scale suitable for assessing sleep quality while remaining attuned to trait-level variations in psychological symptoms. Transcending clinical boundaries enables us to characterise psychiatric risk factors and their relationship with sleep quality, contributing to a deeper understanding of defined psychiatric diagnoses. In line with this transdiagnostic view, we found that COSMOS scores were not only higher for participants with psychiatric diagnoses, with large effect sizes, but also of neuroticism and compulsivity traits, which are psychological risk factors, and recent symptoms of depression. Interestingly there was an overlap in the subscales which contributed most to these associations.

Daytime dysfunction due to sleepiness and bedtime habits such as screen time or reading were top contributors to each of these effects. This finding mirrors what has been reported in the literature about technology use and poor mental health outcomes (Demirci et al., 2015; Elhai et al., 2017). For anxiety and depression, we also observed that the impact of sleep latency and disturbance alongside both of these which is in concordance with symptoms commonly experienced in numerous psychiatric populations (Freeman et al., 2020; Johnson et al., 2006; Mason & Harvey, 2014). Not only does this support associations between our novel scale and psychological traits, but it also demonstrates the contributions of the additional bedtime habit items.

There was no direct corresponding comparison that could be made between the evaluated established scales and our more experimental subscales: bedtime habits and social jetlag. Nonetheless, these subscales were included in COSMOS as they captured information relevant to current sleep behaviour and lifestyle. The bedtime habits scale showed weak correlations with subscales relating to sleep timing and sleep dysfunction. These weak correlations are not entirely surprising as these subscales measure qualities adjacent to that of the established subscales rather than a perfect overlap in content. Social jetlag similarly showed a weak negative correlation to morningness-eveningness but no clear relationship to the other scales. This negative relationship is more surprising as greater differences in weekend and weekday sleep timings have been reported as typical of people with evening-type chronotypes (Roenneberg et al., 2012). It is important to note that several social jetlag calculations have been documented including taking the difference in sleep midpoints or wake times (Roenneberg et al., 2012). Here we calculated the difference in duration which captures differences in sleep length. However, it is possible that a social jetlag calculation that accounts for differences in actual timing or sleep midpoint would link more closely to a chronotype measure.

Both social jetlag and screen time around bedtime have been implicated as risk factors for mental health issues. Moreover, social jetlag has been associated with a greater risk of experiencing depressive symptoms (Levandovski et al., 2011), and sleep quality mediates this relationship between bedtime screen time and poor mental health outcomes (Alonzo et al., 2021). To fully assess the validity and utility of these subscales, it is imperative to evaluate their sensitivity to the measures of interest.

The demonstrated association of COSMOS to psychological traits and its convergence with popular established tools is promising. Our choice of a large, online community is fitting considering the growing emphasis on studying transdiagnostic markers in contemporary psychiatric research (Gillan et al., 2016, 2017). Furthermore, we have shown that COSMOS is suitable for unsupervised online administration, which is essential for researchers moving to large-scale community-based samples.

COSMOS was designed to be comprehensive to address the issue of having to use multiple sleep quality questionnaires to capture different relevant aspects. Although we have achieved this, a consideration is the need for short scales in some studies, where various constructs are collected in the same session. COSMOS is modular, being deployable in relevant sub-sections for different purposes. Future work should focus on exploring whether an abbreviated version, with fewer items per subscale, can be achieved. We identified some items that could be removed or rephrased in future iterations of the scale based on the observed responses and their variability or their sensitivity to other scales. Questions about drug, alcohol, cigarette, and medication quantity did not have the expected spread, with some reported quantities going beyond plausible ranges, albeit these being of low incidence. This highlights the importance of using filters and flags for non-compliant responses. In future versions, asking participants about shiftwork would be helpful when calculating and interpreting certain items such as duration, onset, and social jetlag. Some questions about duration and sleep timing, particularly wake-up time, could be reduced in future versions of the scale due to their sparsity and redundancy.

Although we endeavoured to test a community-based sample, it is important to note that our sample had biases, including toward older, White, and higher educated participants. Even healthy ageing comes with changes in sleep duration, latency, and number of nighttime awakenings (Lavoie et al., 2018; Mander et al., 2017). Similarly, lower socioeconomic status and educational attainment have been related to shorter sleep duration or sleeping too much and more frequent sleep complaints (Grandner et al., 2010; Lee et al., 2021). Our sample was comprised mainly of native English speakers, meaning further validation would be required to ensure the scale’s suitability for use with non-native English speakers. This scale validation study also ran at the end of a longer study which introduces further completion biases. Therefore, generation of normative scores for COSMOS requires further data collection with random sampling. This would also allow for the exploration of the interaction of sleep with clinical labels, psychological traits, or psychiatric symptoms to predict the emergence or intensification of mental health episodes and states.

It is important to note that the data presented here were collected at the beginning of 2022, a period when the effects of the coronavirus disease (COVID-19) pandemic remained significant. COVID-19 infections disrupt sleep and the effects can be prolonged, with recovered patients reporting persistent sleep disturbances and changes in mood and energy levels months after their initial positive test (Nalbandian et al., 2021). Furthermore, environmental changes and disruptions to routine such as continued remote working and isolation orders may have influenced people’s self-reported sleep behaviours (Limongi et al., 2023; Yuan et al., 2022). Finally, the measure of fit in the CFA was moderate for both COSMOS and established scales. This is likely explained by the restrictions imposed on model parameters in CFA, as described by Marsh and colleagues (2014), which increases the likelihood of having moderate model fit even for well-established scales (Marsh et al., 2010).

The assessment of sleep is inherently complex due to its multifaceted nature. Recognising the significance of studying sleep as a transdiagnostic construct in relation to psychiatric disorders is essential as we move towards more diagnostic-label-agnostic methods of understanding, diagnosing, and treating psychiatric disorders. COSMOS presents a new, validated, and sensitive tool optimized for unsupervised administration to enable very large longitudinal studies as mental health research continues to transition to online deployment.

## Data Availability

Data can be made available upon reasonable request to the authors.

## Acknowledgements

We thank the Great British Intelligence Test cohort for their contributions. We also acknowledge Professor Derk-Jan Dijk for his guidance on the initial iterations of the scale.

## Ethical Approval

This project was approved by the Imperial College Research Ethics Committee (17IC4009). Full informed electronic consent was obtained from participants prior to completing this study.

## Consent for Publication

Not applicable

## Data Sharing

Data can be made available upon reasonable request to Professor Adam Hampshire.

## Methodological Statement

We report how we determined our sample size, all data exclusions, all manipulations, and all measures in the study.

## References

1. Akram, U., Stevenson, J. C., Gardani, M., Allen, S., & Johann, A. F. (2023). Personality and insomnia: A systematic review and narrative synthesis. Journal of Sleep Research, 32(6), e14031. 10.1111/jsr.14031

2. Alonzo, R., Hussain, J., Stranges, S., & Anderson, K. K. (2021). Interplay between social media use, sleep quality, and mental health in youth: A systematic review. Sleep Medicine Reviews, 56, 101414. 10.1016/j.smrv.2020.101414

3. Ashton, J. E., & Cairney, S. A. (2021). Future-relevant memories are not selectively strengthened during sleep. PLOS ONE, 16(11), e0258110. 10.1371/journal.pone.0258110

4. Bălăeţ, M., Alhajraf, F., Zerenner, T., Welch, J., Razzaque, J., Lo, C., Giunchiglia, V., Trender, W., Lerede, A., Hellyer, P. J., Manohar, S. G., Malhotra, P., Hu, M., & Hampshire, A. (2024). Online cognitive monitoring technology for people with Parkinson’s disease and REM sleep behavioural disorder. Npj Digital Medicine, 7(1), 1–12. 10.1038/s41746-024-01124-6

5. Biggs, J. (2022). factor-analyzer: A Factor Analysis class (Version 0.4.0) [Computer software]. Educational Testing Service.

6. Binoy, S., Lithwick Algon, A., Ben Adiva, Y., Montaser-Kouhsari, L., & Saban, W. (2024). Online cognitive testing in Parkinson’s disease: Advantages and challenges. Frontiers in Neurology, 15. 10.3389/fneur.2024.1363513

7. Buysse, D. J., Reynolds, C. F., Monk, T. H., Berman, S. R., & Kupfer, D. J. (1989). The Pittsburgh sleep quality index: A new instrument for psychiatric practice and research. Psychiatry Research, 28(2), 193–213. 10.1016/0165-1781(89)90047-4

8. Chamberlain, S. R., & Grant, J. E. (2018). Initial validation of a transdiagnostic compulsivity questionnaire: The Cambridge-Chicago Compulsivity Trait Scale. CNS Spectrums, 23(5), 340–346. 10.1017/S1092852918000810

9. Cronbach, L. J. (1951). Coefficient alpha and the internal structure of tests. Psychometrika, 16(3), 297–334. 10.1007/BF02310555

10. Demirci, K., Akgönül, M., & Akpinar, A. (2015). Relationship of smartphone use severity with sleep quality, depression, and anxiety in university students. Journal of Behavioral Addictions, 4(2), 85–92. 10.1556/2006.4.2015.010

11. Elhai, J. D., Dvorak, R. D., Levine, J. C., & Hall, B. J. (2017). Problematic smartphone use: A conceptual overview and systematic review of relations with anxiety and depression psychopathology. Journal of Affective Disorders, 207, 251–259. 10.1016/j.jad.2016.08.030

12. Ferrarelli, F. (2020). Sleep disturbances in Schizophrenia and Psychosis. Schizophrenia Research, 221, 1–3. 10.1016/j.schres.2020.05.022

13. Foster, R. G. (2020). Sleep, circadian rhythms and health. Interface Focus, 10(3), 20190098. 10.1098/rsfs.2019.0098

14. Freeman, D., Sheaves, B., Waite, F., Harvey, A. G., & Harrison, P. J. (2020). Sleep disturbance and psychiatric disorders. The Lancet Psychiatry, 7(7), 628–637. 10.1016/S2215-0366(20)30136-X

15. Galbiati, A., Verga, L., Giora, E., Zucconi, M., & Ferini-Strambi, L. (2019). The risk of neurodegeneration in REM sleep behavior disorder: A systematic review and meta-analysis of longitudinal studies. Sleep Medicine Reviews, 43, 37–46. 10.1016/j.smrv.2018.09.008

16. Gillan, C. M., Fineberg, N. A., & Robbins, T. W. (2017). A trans-diagnostic perspective on obsessive-compulsive disorder. Psychological Medicine, 47(9), 1528–1548. 10.1017/S0033291716002786

17. Gillan, C. M., Kosinski, M., Whelan, R., Phelps, E. A., & Daw, N. D. (2016). Characterizing a psychiatric symptom dimension related to deficits in goal-directed control. eLife, 5, e11305. 10.7554/eLife.11305

18. Gosling, S. D., Rentfrow, P. J., & Swann, W. B. (2003). A very brief measure of the Big-Five personality domains. Journal of Research in Personality, 37(6), 504–528. 10.1016/S0092-6566(03)00046-1

19. Grandner, M. A., Patel, N. P., Gehrman, P. R., Xie, D., Sha, D., Weaver, T., & Gooneratne, N. (2010). Who gets the best sleep? Ethnic and socioeconomic factors related to sleep complaints. Sleep Medicine, 11(5), 470–478. 10.1016/j.sleep.2009.10.006

20. Gutiérrez, M. L. P., Lugo, J. A. M., Lozano, V. L., & Navarro, D. C. P. (2024). Sleep and Learning: A Systematic Review. International Archives of Otorhinolaryngology. 10.1055/s-0043-1777294

21. Guttesen, A. á V., Gaskell, M. G., Madden, E. V., Appleby, G., Cross, Z. R., & Cairney, S. A. (2023). Sleep loss disrupts the neural signature of successful learning. Cerebral Cortex, 33(5), 1610–1625. 10.1093/cercor/bhac159

22. Hampshire, A. (2020). Great British Intelligence Test Protocol.

23. Hampshire, A., Ballard, C., & Williams, G. (2020). Computerized neuropsychological tests undertaken on digital platforms are cost effective, achieve high engagement, distinguish and are highly sensitive to longitudinal change: Data from the PROTECT and GBIT studies. Alzheimer’s & Dementia, 16(S10), e041122. 10.1002/alz.041122

24. Hampshire, A., Hellyer, P. J., Soreq, E., Mehta, M. A., Ioannidis, K., Trender, W., Grant, J. E., & Chamberlain, S. R. (2021). Associations between dimensions of behaviour, personality traits, and mental-health during the COVID-19 pandemic in the United Kingdom. Nature Communications, 12(1), 4111. 10.1038/s41467-021-24365-5

25. He, S., Hasler, B. P., & Chakravorty, S. (2019). Alcohol and sleep-related problems. Current Opinion in Psychology, 30, 117–122. 10.1016/j.copsyc.2019.03.007

26. Hershner, S. (2020). Sleep and academic performance: Measuring the impact of sleep. Current Opinion in Behavioral Sciences, 33, 51–56. 10.1016/j.cobeha.2019.11.009

27. Horne, J. A., & Ostberg, O. (1976). A self-assessment questionnaire to determine morningness-eveningness in human circadian rhythms. International Journal of Chronobiology, 4(2), 97–110.

28. John, O. P. (1991). The “Big Five” Inventory-versions 4a and 54. Berkeley: University of California, Berkeley, Institute of Personality and Social Research/Institute of Personality and Social Research. https://cir.nii.ac.jp/crid/1370004237628112900

29. Johns, M. W. (1991). A New Method for Measuring Daytime Sleepiness: The Epworth Sleepiness Scale. Sleep, 14(6), 540–545. 10.1093/sleep/14.6.540

30. Johnson, E. O., Roth, T., & Breslau, N. (2006). The association of insomnia with anxiety disorders and depression: Exploration of the direction of risk. Journal of Psychiatric Research, 40(8), 700–708. 10.1016/j.jpsychires.2006.07.008

31. Klingman, K. J., Jungquist, C. R., & Perlis, M. L. (2017). Questionnaires that screen for multiple sleep disorders. Sleep Medicine Reviews, 32, 37–44. 10.1016/j.smrv.2016.02.004

32. Kotov, R., Gamez, W., Schmidt, F., & Watson, D. (2010). Linking “big” personality traits to anxiety, depressive, and substance use disorders: A meta-analysis. Psychological Bulletin, 136(5), 768–821. 10.1037/a0020327

33. Kroenke, K., Spitzer, R. L., & Williams, J. B. W. (2001). The PHQ-9. Journal of General Internal Medicine, 16(9), 606–613. 10.1046/j.1525-1497.2001.016009606.x

34. Kucharczyk, E. R., Morgan, K., & Hall, A. P. (2012). The occupational impact of sleep quality and insomnia symptoms. Sleep Medicine Reviews, 16(6), 547–559. 10.1016/j.smrv.2012.01.005

35. Lahey, B. B. (2009). Public Health Significance of Neuroticism. American Psychologist, 64(4), 241–256. 10.1037/a0015309

36. Lavoie, C. J., Zeidler, M. R., & Martin, J. L. (2018). Sleep and aging. Sleep Science and Practice, 2(1), 3. 10.1186/s41606-018-0021-3

37. Lee, G. B., Kim, H. C., Jeon, Y. J., & Jung, S. J. (2021). Association between socioeconomic status and longitudinal sleep quality patterns mediated by depressive symptoms. Sleep, 44(8), zsab044. 10.1093/sleep/zsab044

38. Levandovski, R., Dantas, G., Fernandes, L. C., Caumo, W., Torres, I., Roenneberg, T., Hidalgo, M. P. L., & Allebrandt, K. V. (2011). Depression Scores Associate With Chronotype and Social Jetlag in a Rural Population. Chronobiology International, 28(9), 771–778. 10.3109/07420528.2011.602445

39. Limongi, F., Siviero, P., Trevisan, C., Noale, M., Catalani, F., Ceolin, C., Conti, S., di Rosa, E., Perdixi, E., Remelli, F., Prinelli, F., & Maggi, S. (2023). Changes in sleep quality and sleep disturbances in the general population from before to during the COVID-19 lockdown: A systematic review and meta-analysis. Frontiers in Psychiatry, 14. 10.3389/fpsyt.2023.1166815

40. Liu, S., Wing, Y. K., Hao, Y., Li, W., Zhang, J., & Zhang, B. (2019). The associations of long-time mobile phone use with sleep disturbances and mental distress in technical college students: A prospective cohort study. Sleep, 42(2). 10.1093/sleep/zsy213

41. Mander, B. A., Winer, J. R., & Walker, M. P. (2017). Sleep and Human Aging. Neuron, 94(1), 19–36. 10.1016/j.neuron.2017.02.004

42. Marsh, H. W., Morin, A. J. S., Parker, P. D., & Kaur, G. (2014). Exploratory Structural Equation Modeling: An Integration of the Best Features of Exploratory and Confirmatory Factor Analysis. Annual Review of Clinical Psychology, 10(1), 85–110. 10.1146/annurev-clinpsy-032813-153700

43. Mason, E. C., & Harvey, A. G. (2014). Insomnia before and after treatment for anxiety and depression. Journal of Affective Disorders, 168, 415–421. 10.1016/j.jad.2014.07.020

44. Morton, E., Hou, S. H., Fogarty, O., Murray, G., Barnes, S., Depp, C., Crest, BD, & Michalak, E. (2020). A Web-Based Adaptation of the Quality of Life in Bipolar Disorder Questionnaire: Psychometric Evaluation Study. JMIR Mental Health, 7(4), e17497. 10.2196/17497

45. Nalbandian, A., Sehgal, K., Gupta, A., Madhavan, M. V., McGroder, C., Stevens, J. S., Cook, J. R., Nordvig, A. S., Shalev, D., Sehrawat, T. S., Ahluwalia, N., Bikdeli, B., Dietz, D., Der-Nigoghossian, C., Liyanage-Don, N., Rosner, G. F., Bernstein, E. J., Mohan, S., Beckley, A. A., … Wan, E. Y. (2021). Post-acute COVID-19 syndrome. Nature Medicine, 27(4), 601–615. 10.1038/s41591-021-01283-z

46. Ormel, J., Jeronimus, B. F., Kotov, R., Riese, H., Bos, E. H., Hankin, B., Rosmalen, J. G. M., & Oldehinkel, A. J. (2013). Neuroticism and common mental disorders: Meaning and utility of a complex relationship. Clinical Psychology Review, 33(5), 686–697. 10.1016/j.cpr.2013.04.003

47. R Core Team. (2022). R: A Language and Environment for Statistical Computing (Version 4.2.2 (2022-10-31)) [Computer software]. R Foundation for Statistical Computing. https://www.R-project.org/

48. Ramar, K., Malhotra, R. K., Carden, K. A., Martin, J. L., Abbasi, -Feinberg Fariha, Aurora, R. N., Kapur, V. K., Olson, E. J., Rosen, C. L., Rowley, J. A., Shelgikar, A. V., & Trotti, L. M. (2021). Sleep is essential to health: An American Academy of Sleep Medicine position statement. Journal of Clinical Sleep Medicine, 17(10), 2115–2119. 10.5664/jcsm.9476

49. Roenneberg, T., Allebrandt, K. V., Merrow, M., & Vetter, C. (2012). Social jetlag and obesity. Current Biology, 22(10), 939–943. 10.1016/J.CUB.2012.03.038/ATTACHMENT/DC17B376-110B-40FC-8B82-D81509CD2436/MMC1.PDF

50. Rosseel, Y. (2012). lavaan: An R Package for Structural Equation Modeling. 48(2), 1–36. 10.18637/jss.v048.i02

51. Schirmbeck, F., Boyette, L.-L., Valk, R. van der, Meijer, C., Dingemans, P., Van, R., de Haan, L., Kahn, R. S., de Haan, L., van Os, J., Wiersma, D., Bruggeman, R., Cahn, W., Meijer, C., & Myin-Germeys, I. (2015). Relevance of Five-Factor Model personality traits for obsessive–compulsive symptoms in patients with psychotic disorders and their un-affected siblings. Psychiatry Research, 225(3), 464–470. 10.1016/j.psychres.2014.11.066

52. Scott, H., & Woods, H. C. (2019). Understanding Links Between Social Media Use, Sleep and Mental Health: Recent Progress and Current Challenges. Current Sleep Medicine Reports, 5(3), 141–149. 10.1007/s40675-019-00148-9

53. Seabold, S., & Perktold, J. (2010). Statsmodels: Econometric and Statistical Modeling with Python. 92–96. 10.25080/Majora-92bf1922-011

54. Segalàs, C., Labad, J., Salvat-Pujol, N., Real, E., Alonso, P., Bertolín, S., Jiménez-Murcia, S., Soriano-Mas, C., Monasterio, C., Menchón, J. M., & Soria, V. (2021). Sleep disturbances in obsessive-compulsive disorder: Influence of depression symptoms and trait anxiety. BMC Psychiatry, 21(1), 42. 10.1186/s12888-021-03038-z

55. Spitzer, R. L., Kroenke, K., Williams, J. B. W., & Löwe, B. (2006). A brief measure for assessing generalized anxiety disorder: The GAD-7. Archives of Internal Medicine, 166(10), 1092–1097. 10.1001/archinte.166.10.1092

56. The Pandas Development Team. (2022). pandas-dev/pandas: Pandas (Version 1.4.4) [Computer software]. Zenodo. 10.5281/zenodo.3509134

57. Tiego, J., Trender, W., Hellyer, P. J., Grant, J. E., Hampshire, A., & Chamberlain, S. R. (2023). Measuring Compulsivity as a Self-Reported Multidimensional Transdiagnostic Construct: Large-Scale (N = 182,000) Validation of the Cambridge–Chicago Compulsivity Trait Scale. Assessment, 30(8), 2433–2448. 10.1177/10731911221149083

58. Vallat, R. (2018). Pingouin: Statistics in Python. Journal of Open Source Software, 3(31), 1026. 10.21105/joss.01026

59. Virtanen, P., Gommers, R., Oliphant, T. E., Haberland, M., Reddy, T., Cournapeau, D., Burovski, E., Peterson, P., Weckesser, W., Bright, J., van der Walt, S. J., Brett, M., Wilson, J., Millman, K. J., Mayorov, N., Nelson, A. R. J., Jones, E., Kern, R., Larson, E., … van Mulbregt, P. (2020). SciPy 1.0: Fundamental algorithms for scientific computing in Python. Nature Methods, 17(3), Article 3. 10.1038/s41592-019-0686-2

60. Waite, F., Sheaves, B., Isham, L., Reeve, S., & Freeman, D. (2020). Sleep and schizophrenia: From epiphenomenon to treatable causal target. Schizophrenia Research, 221, 44–56. 10.1016/j.schres.2019.11.014

61. Watson, N. F., Badr, M. S., Belenky, G., Bliwise, D. L., Buxton, O. M., Buysse, D., Dinges, D. F., Gangwisch, J., Grandner, M. A., Kushida, C., Malhotra, R. K., Martin, J. L., Patel, S. R., Quan, S. F., & Tasali, E. (2015). Joint Consensus Statement of the American Academy of Sleep Medicine and Sleep Research Society on the Recommended Amount of Sleep for a Healthy Adult: Methodology and Discussion. Journal of Clinical Sleep Medicine, 11(08), 931–952. 10.5664/jcsm.4950

62. Yuan, R. K., Zitting, K.-M., Maskati, L., & Huang, J. (2022). Increased sleep duration and delayed sleep timing during the COVID-19 pandemic. Scientific Reports, 12(1), 10937. 10.1038/s41598-022-14782-x

