## Supplemental Materials for "Validation of the Comprehensive Online Sleep Monitoring Scale (COSMOS) in a Large Population Sample"

### Appendix A: Sleep Scales Utilised in COSMOS Development

|  | Questionnaire | Acronym |
| --- | --- | --- |
| 1 | Holland Sleep Disorders Questionnaire | HSDQ |
| 2 | Medical Outcomes Study Sleep Scale | MOS-SS |
| 3 | Auckland Sleep Questionnaire | ASQ |
| 4 | Basic Nordic Sleep Questionnaire | BNSQ |
| 5 | Sleep Disorders Questionnaire | SDQ |
| 6 | Pittsburgh Sleep Quality Index | PSQI |
| 7 | SLEEP-50 Questionnaire | SLEEP-50 |
| 8 | Iowa Sleep Disturbances Index | ISDI |
| 9 | Leeds Sleep Evaluation Questionnaire | LSEQ |
| 10 | Groningen Sleep Quality Scale | GSQS |
| 11 | General Sleep Disturbance Scale | GSDS |
| 12 | Jenkins Sleep Scale | JSS |
| 13 | Epworth Sleepiness Scale | ESS |

**References**

Arroll, B., Fernando, A., 3rd, Falloon, K., Warman, G., & Goodyear-Smith, F. (2011). Development, validation (diagnostic accuracy) and audit of the Auckland Sleep Questionnaire: a new tool for diagnosing causes of sleep disorders in primary care. *Journal of primary health care*, *3*(2), 107–113.

Buysse, D. J., Reynolds, C. F., Monk, T. H., Berman, S. R., & Kupfer, D. J. (1989). The Pittsburgh Sleep Quality Index: A new instrument for psychiatric practice and research. *Psychiatry Research, 28*(2), 193-213. <https://doi.org/10.1016/0165-1781(89)90047-4>

Douglass, A. B., Bornstein, R., Nino-Murcia, G., Keenan, S., Miles, L., Zarcone, V. P., Jr, Guilleminault, C., & Dement, W. C. (1994). The Sleep Disorders Questionnaire. I: Creation and multivariate structure of SDQ. *Sleep*, *17*(2), 160–167. <https://doi.org/10.1093/sleep/17.2.160>

Hays, R. D., & Stewart, A. L. (1992). Sleep measures. In A. L. Stewart & J. E. Ware (eds.), Measuring functioning and well-being: The Medical Outcomes Study approach (pp. 235-259), Durham, NC: Duke University Press.

Johns, M. W. (1991). A new method for measuring daytime sleepiness: The Epworth Sleepiness Scale. *Sleep, 14*(6), 540-545. <https://doi.org/10.1093/sleep/14.6.540>

Kerkhof, G. A., Geuke, M. E., Brouwer, A., Rijsman, R. M., Schimsheimer, R. J., & Van Kasteel, V. (2013). Holland Sleep Disorders Questionnaire: a new sleep disorders questionnaire based on the International Classification of Sleep Disorders-2. *Journal of sleep research*, *22*(1), 104–107. https://doi.org/10.1111/j.1365-2869.2012.01041.x

Koffel E, Watson D. Development and initial validation of the Iowa Sleep Disturbances Inventory. Assessment. 2010 Dec;17(4):423-39. doi: 10.1177/1073191110362864.

Lee K. A. (1992). Self-reported sleep disturbances in employed women. *Sleep*, *15*(6), 493–498. <https://doi.org/10.1093/sleep/15.6.493>

Meijman, T. F., de Vries-Griever, A. H., De Vries, G., & Kampman, R. (1988). The evaluation of the Groningen sleep quality scale. *Groningen: Heymans Bulletin (HB 88-13-EX)*, *2006*.

Parrott, A. C., & Hindmarch, I. (1980). The Leeds Sleep Evaluation Questionnaire in psychopharmacological investigations - a review. *Psychopharmacology*, *71*(2), 173–179. <https://doi.org/10.1007/BF00434408>

Partinen, M., & Gislason, T. (1995). Basic Nordic Sleep Questionnaire (BNSQ): a quantitated measure of subjective sleep complaints. *Journal of sleep research*, *4*(S1), 150–155. <https://doi.org/10.1111/j.1365-2869.1995.tb00205.x>

Shahid, A., Wilkinson, K., Marcu, S., & Shapiro, C. M. (2011). Jenkins sleep scale. In *Stop, that and one hundred other sleep scales* (pp. 203-204). Springer, New York, NY.

Spoormaker, V. I., Verbeek, I., van den Bout, J., & Klip, E. C. (2005). Initial validation of the SLEEP-50 questionnaire. *Behavioral sleep medicine*, *3*(4), 227–246. https://doi.org/10.1207/s15402010bsm0304_4

### Appendix B: CHI-T EFA Results

Based on the scree plot and previous literature suggesting a two-factor solution (Chamberlain & Grant, 2018; Tiego et al., 2023), an exploratory factor analysis using the principal axis factoring method with varimax rotation was applied on 5237 cases to extract two factors. Bartlett’s test of sphericity confirmed the correlation matrix was not random (*χ^2^* (105) = 18833.53 *p* < .001) and the Kaiser-Meyer Olkin (KMO) statistic was 0.87, well above the minimum threshold of 0.50 for conducting exploratory factor analysis. The factor structure was clear and interpretable, with most items loading onto the factors in the same pattern reported by (Tiego et al., 2023; **Table 1**).

The eigenvalues and percentage of variance explained by each factor are shown in **Table 2**.

| Table 1 |  | |  |
| --- | --- | --- | --- |
| *Factor Loadings and Communalities for Varimax Rotated Two-Factor Solution for 15 CHI-T Items (N = 5237)* | | | |
|  | Factor Loading | |  |
|  | 1 | 2 | Communality |
| Need for completion | **0.53** | -0.14 | 0.30 |
| Doing things just right | **0.72** | -0.02 | 0.52 |
| Repetition to meet high standard | **0.71** | 0.07 | 0.50 |
| Getting stuck in thoughts | 0.39 | **0.50** | 0.40 |
| Habit propensity | **0.35** | 0.23 | 0.17 |
| Addictive propensity | 0.14 | **0.59** | 0.36 |
| Rigidity | 0.24 | **0.40** | 0.22 |
| Tendency to act of urges | -0.03 | **0.56** | 0.32 |
| Doing things that are immediately rewarding | -0.08 | **0.66** | 0.44 |
| Difficulty moving from task to task | **0.63** | 0.30 | 0.49 |
| High standards | **0.59** | 0.09 | 0.36 |
| Completion leads to soothing | **0.63** | 0.17 | 0.42 |
| Need for control | **0.35** | 0.31 | 0.21 |
| Needing to be the best | **0.44** | 0.34 | 0.31 |
| Scope for improvement/nothing is good enough | **0.15** | 0.08 | 0.03 |
| *Factors correspond to 1) Perfectionism and 2) Compulsive Soothing.* | | | |

**Figure 1**

Scree Plot of Observed Eigenvalues

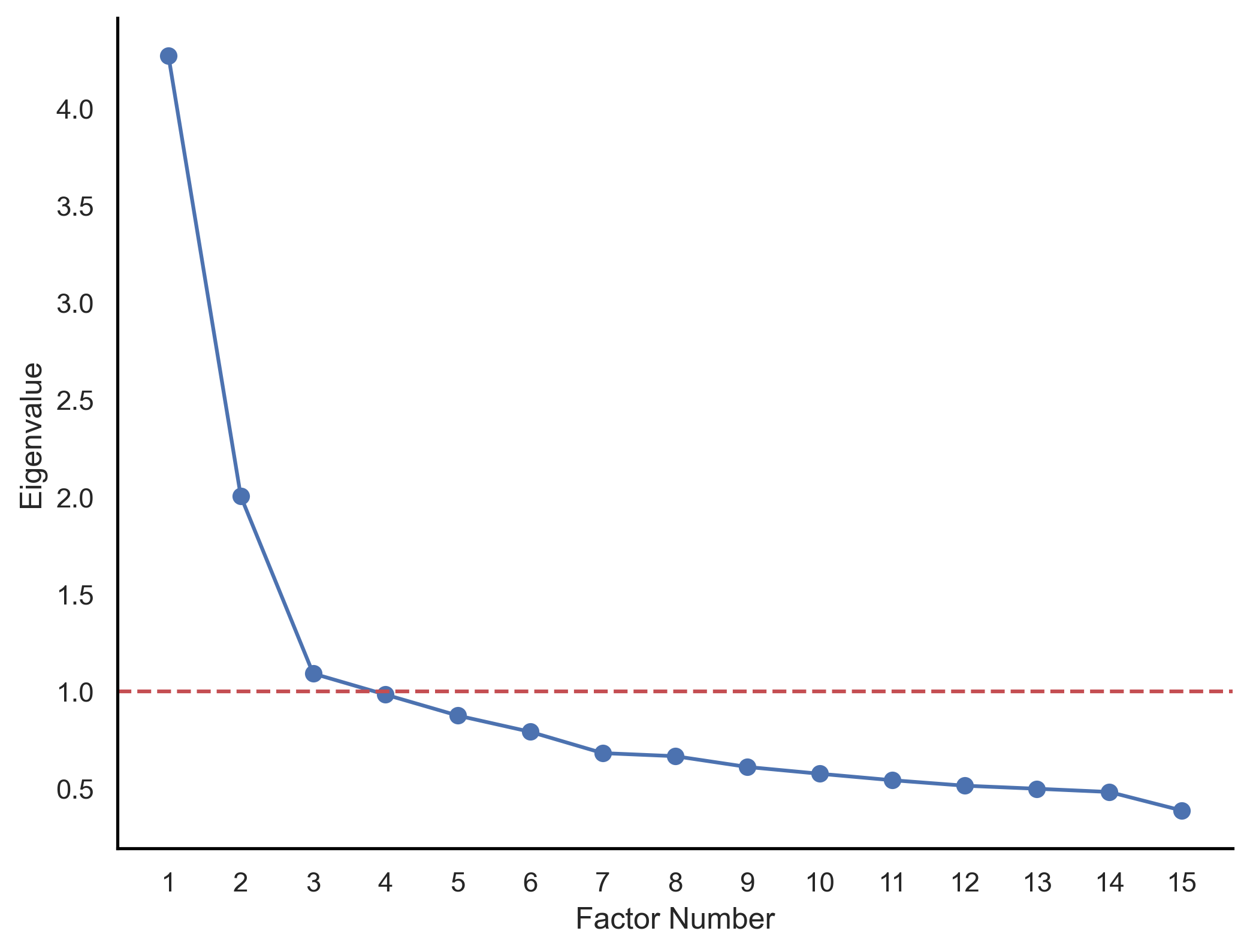

| Table 2 |  |  |  |
| --- | --- | --- | --- |
| *Eigenvalues, Percentages of Variance, and Cumulative Percentages for Factors for 15 CHI-T Items* | | | |
| Factor | Eigenvalue | % of variance | Cumulative % |
| 1 | 4.27 | 20.89 | 20.89 |
| 2 | 2.01 | 12.76 | 33.65 |

**References**

Chamberlain, S. R., & Grant, J. E. (2018). Initial validation of a transdiagnostic compulsivity questionnaire: The Cambridge-Chicago Compulsivity Trait Scale. *CNS Spectrums*, *23*(5), 340–346. <https://doi.org/10.1017/S1092852918000810>

Tiego, J., Trender, W., Hellyer, P. J., Grant, J. E., Hampshire, A., & Chamberlain, S. R. (2023). Measuring Compulsivity as a Self-Reported Multidimensional Transdiagnostic Construct: Large-Scale (N = 182,000) Validation of the Cambridge–Chicago Compulsivity Trait Scale. *Assessment*, *30*(8), 2433–2448. https://doi.org/10.1177/10731911221149083

### Appendix C: Big Five EFA Results

An exploratory factor analysis was applied to 5236 cases of the Big Five short questionnaire items using the principal axis method of extraction and varimax rotation. Five factors were extracted after considering the scree plot and existing literature. Bartlett’s test of sphericity confirmed the correlation matrix was not random (*χ^2^* (153) = 31336.86 *p* < .001) and the Kaiser-Meyer Olkin (KMO) statistic was 0.74 above the minimum threshold, indicating the data was suitable for exploratory factor analysis. The factor structure was clear and interpretable, with items for each established personality factor loading onto the expected factors (Gosling et al., 2003; John, 1991; **Table 1**).

The eigenvalues and variance explained by the five factors are summarized in **Table 2**.

| **Table 1** |  |  |  |  |  |  |
| --- | --- | --- | --- | --- | --- | --- |
| *Factor Loadings and Communalities for Varimax Rotated Five-Factor Solution for 18 Big Five Items (N = 5236)* | | | | | | |
|  | Factor Loading | | | | |  |
|  | 1 | 2 | 3 | 4 | 5 | Communality |
| Inventive | **0.80** | -0.10 | 0.18 | -0.12 | 0.03 | 0.70 |
| Original | **0.79** | -0.17 | 0.14 | -0.11 | 0.07 | 0.70 |
| Reflect / Play with Ideas | **0.55** | 0.00 | 0.05 | 0.20 | 0.01 | 0.34 |
| Artistic | **0.55** | -0.02 | -0.17 | 0.47 | -0.02 | 0.55 |
| Sophisticated in the arts | **0.44** | -0.02 | -0.13 | 0.39 | -0.01 | 0.36 |
| Perseveres | 0.13 | 0.04 | 0.06 | 0.06 | **0.70** | 0.51 |
| Disorganised | 0.12 | 0.01 | -0.09 | -0.01 | **-0.48** | 0.26 |
| Talkative | 0.10 | **-0.74** | -0.05 | 0.14 | 0.00 | 0.58 |
| Considerate and Kind | 0.09 | -0.06 | 0.08 | **0.52** | 0.11 | 0.30 |
| Thorough | 0.08 | -0.03 | 0.01 | 0.07 | **0.71** | 0.52 |
| Relaxed | 0.07 | -0.01 | **0.80** | 0.15 | 0.06 | 0.67 |
| Forgiving | 0.03 | 0.01 | 0.25 | **0.44** | -0.04 | 0.26 |
| Emotionally Stable | 0.01 | 0.05 | **0.74** | 0.16 | 0.13 | 0.59 |
| Cooperative | 0.00 | -0.15 | 0.14 | **0.42** | 0.09 | 0.23 |
| Worrier | -0.03 | 0.08 | **-0.68** | 0.00 | -0.03 | 0.48 |
| Quiet | -0.05 | **0.92** | -0.03 | -0.03 | 0.00 | 0.84 |
| Reserved | -0.07 | **0.79** | -0.07 | -0.06 | 0.00 | 0.64 |
| Few Artistic Interests | **-0.44** | 0.01 | 0.16 | -0.35 | 0.03 | 0.34 |
| *Factors correspond to 1) Openness, 2) Extraversion (reversed), 3) Neuroticism (reversed), 4) Agreeableness, and 5) Conscientiousness.* | | | | | | |

**Figure 1**

Scree Plot of Observed Eigenvalues

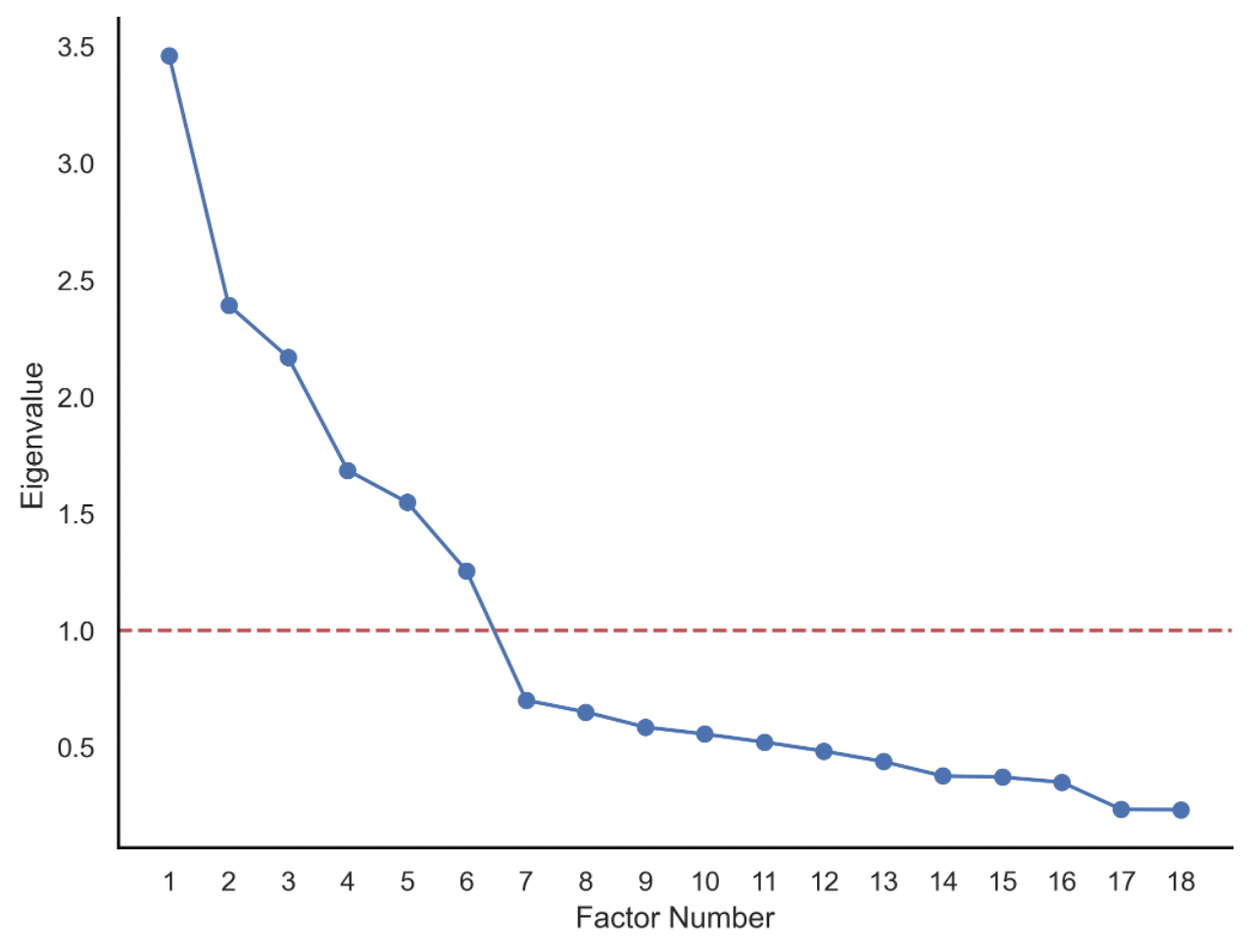

| **Table 2** |  |  |  |
| --- | --- | --- | --- |
| *Eigenvalues, Percentages of Variance, and Cumulative Percentages for Factors for 18 Big Five Items* | | | |
| Factor | Eigenvalue | % of variance | Cumulative % |
| 1 | 3.46 | 12.94 | 12.94 |
| 2 | 2.39 | 11.64 | 24.58 |
| 3 | 2.17 | 10.46 | 35.04 |
| 4 | 1.68 | 7.13 | 42.18 |
| 5 | 1.55 | 7.13 | 49.30 |

**References**

Gosling, S. D., Rentfrow, P. J., & Swann, W. B. (2003). A very brief measure of the Big-Five personality domains. *Journal of Research in Personality*, *37*(6), 504–528. <https://doi.org/10.1016/S0092-6566(03)00046-1>

John, O. P. (1991). The “Big Five” Inventory-versions 4a and 54. *Berkeley: University of California, Berkeley, Institute of Personality and Social Research/Institute of Personality and Social Research*. https://cir.nii.ac.jp/crid/1370004237628112900

### Appendix D: COSMOS Eight-Factor Model Loadings

|  | Estimate | Std. Err. | z | *p* |
| --- | --- | --- | --- | --- |
| **Disturbance** |  |  |  |  |
| 1. Find it difficult to fall asleep at night… | 0.59 | 0.01 | 46.79 | <.001 |
| 2. Find it difficult to stay asleep at night… | 0.82 | 0.01 | 73.19 | <.001 |
| 3. Find it difficult to fall asleep because your… | 0.59 | 0.01 | 46.31 | <.001 |
| 4. Awaken with racing thoughts or worries… | 0.62 | 0.01 | 49.63 | <.001 |
| 5. Have trouble sleeping at night due to your… | 0.32 | 0.01 | 23.56 | <.001 |
| 6. Have trouble sleeping at night due to your… | 0.29 | 0.01 | 21.08 | <0.001 |
| 9. Frequently wake up during the night… | 0.72 | 0.01 | 60.2 | <.001 |
| 10. Find it difficult to fall asleep after waking… | 0.79 | 0.01 | 69.6 | <.001 |
| 11. Wake up earlier than planned and were not able… | 0.63 | 0.01 | 51.06 | <.001 |
| 21. Feel that your sleep was disturbed due to mov… | 0.39 | 0.01 | 29.23 | <.001 |
| 64. I have a medical condition that makes it diff… | 0.24 | 0.01 | 17.67 | <.001 |
| 63. I share a bed or room.… | -0.01 | 0.01 | -0.82 | .412 |
| 61. I normally wake up [X] times during the night | 0.64 | 0.01 | 51.96 | <.001 |
| **Dysfunction** |  |  |  |  |
| 15. Feel tired and fatigued during the day | 0.72 | 0.01 | 58.54 | <.001 |
| 16. Take naps during the day | 0.4 | 0.01 | 29.26 | <.001 |
| 17. Were your naps scheduled? | 0.24 | 0.01 | 16.93 | <.001 |
| 18. What was your average nap duration? | -0.17 | 0.01 | -11.98 | <.001 |
| 19. Find it difficult to fulfil work and everyday… | 0.82 | 0.01 | 69.88 | <.001 |
| 20. Feel sleepy during the day which made it… | 0.8 | 0.01 | 67.58 | <.001 |
| **Disorders** |  |  |  |  |
| 22. Sleepwalk | 0.35 | 0.02 | 22.23 | <.001 |
| 23. Act out your dreams | 0.42 | 0.02 | 26.7 | <.001 |
| 24. Did this disturb your partner / members of… | 0.38 | 0.02 | 24.12 | <.001 |
| 25. Grind your teeth or clench your jaw during… | 0.36 | 0.02 | 24.09 | <.001 |
| 26. Have leg twitching or jerking during sleep | 0.49 | 0.01 | 33.3 | <.001 |
| 27. Have trouble sleeping or wake up at night… | 0.56 | 0.01 | 38.57 | <.001 |
| 7. Have trouble sleeping at night due to your… | 0.4 | 0.01 | 26.97 | <.001 |
| 8. Have trouble sleeping due to other discomfort… | 0.52 | 0.02 | 34.04 | <.001 |
| **Bedtime Habits** |  |  |  |  |
| 28. Use a phone or laptop before going to sleep… | 0.69 | 0.01 | 58.16 | <.001 |
| 29. Did you use a dark screen / night mode? | 0.46 | 0.01 | 35.3 | <.001 |
| 30. Feel compelled to check your email, social media… | 0.92 | 0.01 | 87.52 | <.001 |
| 31. Did you try to stop yourself but failed? | 0.64 | 0.01 | 50.85 | <.001 |
| 32. Did you check work related content… | 0.63 | 0.01 | 49.68 | <.001 |
| 33. Did you check social media… | 0.87 | 0.01 | 81.77 | <.001 |
| 34. Watch videos before going to sleep… | 0.42 | 0.01 | 31.91 | <.001 |
| 35. Read before going to sleep… | 0.09 | 0.01 | 6.93 | <.001 |
| 36. Did you read physical or electronic copies… | 0.12 | 0.01 | 9 | <.001 |
| 38. Give up trying to sleep and read instead | 0.16 | 0.01 | 11.8 | <.001 |
| 39. Give up trying to sleep and used your phone… | 0.52 | 0.01 | 40.46 | <.001 |
| 40. Meditate or pray before going to sleep | -0.01 | 0.01 | -0.9 | .370 |
| **Medication** |  |  |  |  |
| 41. Drink alcohol in the evening | 0.44 | 0.02 | 24.12 | <.001 |
| 43. Smoke cigarettes | 0.26 | 0.02 | 14.13 | <.001 |
| 45. Take prescription drugs | 0.56 | 0.02 | 28.62 | <.001 |
| 47. Take over the counter or herbal drugs | 0.44 | 0.02 | 24.23 | <.001 |
| 49. Take illegal drugs | 0.19 | 0.02 | 10.44 | <.001 |
| **Latency** |  |  |  |  |
| 51. I go to bed at… | -0.01 | 0.01 | -0.49 | 0.626 |
| 52. Once I’m in bed, I start trying to fall asleep… | -0.08 | 0.01 | -5.69 | <.001 |
| 53. Once I decide to sleep, falling asleep… | -0.1 | 0.01 | -7.51 | <.001 |
| 54. I sleep for.… | 0.06 | 0.01 | 4.5 | <.001 |
| 55. My sleep schedule differs from weekdays to… | -0.98 | 0.01 | -91.57 | <.001 |
| 56a. On weekdays, I go to bed at… | -0.75 | 0.01 | -65.52 | <.001 |
| 56b. On weekdays, I wake up at… | -0.92 | 0.01 | -85.76 | <.001 |
| 57a. On weekends, I go to bed at… | -0.62 | 0.01 | -50.68 | <.001 |
| 57b. On weekends, I wake up at… | -0.94 | 0.01 | -89.29 | <.001 |
| 58. I wake up at… | -0.85 | 0.01 | -76.2 | <.001 |
| 59. For me, a good night’s sleep lasts… | -0.1 | 0.01 | -7.38 | <.001 |
| 12. Go to sleep and wake up at the same time… | 0.51 | 0.08 | 6.04 | <.001 |
| **Chronotype** |  |  |  |  |
| 60. I naturally prefer to sleep and wake up early… | 0.33 | 0.02 | 18.03 | <.001 |
| 55. My sleep schedule differs from weekdays to… | 0.02 | 0 | 4.07 | <.001 |
| 12. Go to sleep and wake up at the same time… | -0.97 | 0.08 | -11.64 | <.001 |
| **Quality** |  |  |  |  |
| 62. I would rate the quality of my sleep as… | 0.86 | 0.01 | 74.31 | <.001 |
| 13. Have very deep sleep… | 0.63 | 0.01 | 49.41 | <.001 |
| 14. Wake up feeling energized and rested… | 0.66 | 0.01 | 52.7 | <.001 |

### Appendix E: COSMOS Eleven-Factor Model Loadings

|  | Estimate | Std. Err. | z | *p* |
| --- | --- | --- | --- | --- |
| **Disturbance** | |  |  |  |
| 2. Find it difficult to stay asleep at night… | 0.83 | 0.01 | 74.49 | <.001 |
| 3. Find it difficult to fall asleep because your… | 0.56 | 0.01 | 43.94 | <.001 |
| 4. Awaken with racing thoughts or worries… | 0.61 | 0.01 | 48.69 | <.001 |
| 5. Have trouble sleeping at night due to your… | 0.31 | 0.01 | 22.65 | <.001 |
| 6. Have trouble sleeping at night due to your… | 0.27 | 0.01 | 20.17 | <.001 |
| 9. Frequently wake up during the night… | 0.73 | 0.01 | 61.96 | <.001 |
| 10. Find it difficult to fall asleep after waking… | 0.8 | 0.01 | 70.66 | <.001 |
| 11. Wake up earlier than planned and were not able… | 0.64 | 0.01 | 52.1 | <.001 |
| 21. Feel that your sleep was disturbed due to mov… | 0.38 | 0.01 | 28.72 | <.001 |
| 64. I have a medical condition that makes it diff… | 0.24 | 0.01 | 17.36 | <.001 |
| 63. I share a bed or room.… | 0 | 0.01 | -0.19 | .850 |
| 61. I normally wake up [X] times during the night | 0.65 | 0.01 | 53.04 | <.001 |
| **Dysfunction** | |  |  |  |
| 15. Feel tired and fatigued during the day | 0.72 | 0.01 | 58.37 | <.001 |
| 16. Take naps during the day | 0.4 | 0.01 | 29.26 | <.001 |
| 17. Were your naps scheduled? | 0.24 | 0.01 | 16.93 | <.001 |
| 18. What was your average nap duration? | -0.17 | 0.01 | -11.9 | <.001 |
| 19. Find it difficult to fulfil work and everyday… | 0.82 | 0.01 | 70.21 | <.001 |
| 20. Feel sleepy during the day which made it… | 0.8 | 0.01 | 67.61 | <.001 |
| **Disorders** | |  |  |  |
| 22. Sleepwalk | 0.34 | 0.02 | 22.11 | <.001 |
| 23. Act out your dreams | 0.42 | 0.02 | 26.67 | <.001 |
| 24. Did this disturb your partner / members of… | 0.37 | 0.02 | 24.07 | <.001 |
| 25. Grind your teeth or clench your jaw during… | 0.36 | 0.02 | 24.13 | <.001 |
| 26. Have leg twitching or jerking during sleep | 0.49 | 0.01 | 33.36 | <.001 |
| 27. Have trouble sleeping or wake up at night… | 0.56 | 0.01 | 38.61 | <.001 |
| 7. Have trouble sleeping at night due to your… | 0.4 | 0.01 | 26.89 | <.001 |
| 8. Have trouble sleeping due to other discomfort… | 0.51 | 0.02 | 33.98 | <.001 |
| **Bedtime Habits** | |  |  |  |
| 28. Use a phone or laptop before going to sleep… | 0.69 | 0.01 | 58.29 | <.001 |
| 29. Did you use a dark screen / night mode? | 0.46 | 0.01 | 35.39 | <.001 |
| 30. Feel compelled to check your email, social media… | 0.91 | 0.01 | 87.35 | <.001 |
| 31. Did you try to stop yourself but failed? | 0.64 | 0.01 | 50.88 | <.001 |
| 32. Did you check work related content… | 0.63 | 0.01 | 49.59 | <.001 |
| 33. Did you check social media… | 0.87 | 0.01 | 81.72 | <.001 |
| 34. Watch videos before going to sleep… | 0.42 | 0.01 | 32.01 | <.001 |
| 35. Read before going to sleep… | 0.1 | 0.01 | 6.96 | <.001 |
| 36. Did you read physical or electronic copies… | 0.12 | 0.01 | 9.03 | <.001 |
| 38. Give up trying to sleep and read instead | 0.16 | 0.01 | 11.9 | <.001 |
| 39. Give up trying to sleep and used your phone… | 0.52 | 0.01 | 40.61 | <.001 |
| 40. Meditate or pray before going to sleep | -0.01 | 0.01 | -0.9 | .370 |
| **Medication** | |  |  |  |
| 41. Drink alcohol in the evening | 0.45 | 0.02 | 24.07 | <.001 |
| 43. Smoke cigarettes | 0.27 | 0.02 | 14.22 | <.001 |
| 45. Take prescription drugs | 0.56 | 0.02 | 28.44 | <.001 |
| 47. Take over the counter or herbal drugs | 0.44 | 0.02 | 24.04 | <.001 |
| 49. Take illegal drugs | 0.2 | 0.02 | 10.55 | <.001 |
| **Latency** |  |  |  |  |
| 53. Once I decide to sleep, falling asleep… | 0.53 | 0.01 | 38.09 | <.001 |
| 1. Find it difficult to fall asleep at night… | 0.91 | 0.02 | 59.04 | <.001 |
| **Duration** | |  |  |  |
| 54. I sleep for.… | 1.31 | 0.06 | 22.2 | <.001 |
| 59. For me, a good night’s sleep lasts… | 0.38 | 0.02 | 17.99 | <.001 |
| **Sleep Onset** | |  |  |  |
| 51. I go to bed at | 1 | 0.01 | 107.84 | <.001 |
| **Social Jetlag** | |  |  |  |
| Calculated using 56 & 57 | 0.13 | 0.02 | 5.49 | <.001 |
| 55. My sleep schedule differs from weekdays to… (reversed) | 0.89 | 0.14 | 6.57 | <.001 |
| **Chronotype** | |  |  |  |
| 60. I naturally prefer to sleep and wake up early… | 0.46 | 0.02 | 22.07 | <.001 |
| 12. Go to sleep and wake up at the same time… | -0.46 | 0.02 | -22.09 | <.001 |
| **Quality** |  |  |  |  |
| 62. I would rate the quality of my sleep as… | 0.87 | 0.01 | 76.31 | <.001 |
| 13. Have very deep sleep… | 0.62 | 0.01 | 48.94 | <.001 |
| 14. Wake up feeling energized and rested… | 0.66 | 0.01 | 52.17 | <.001 |

### Appendix F: Established Scales Factor Model Loadings

|  | Estimate | Std. Err. | z | *p* |
| --- | --- | --- | --- | --- |
| **DISTURBANCE** | |  |  |  |
| PSQI 5b | 0.71 | 0.01 | 51.66 | <.001 |
| PSQI 5c | 0.39 | 0.01 | 26.25 | <.001 |
| PSQI 5d | 0.33 | 0.02 | 21.8 | <.001 |
| PSQI 5e | 0.28 | 0.02 | 18.62 | <.001 |
| PSQI 5f | 0.29 | 0.01 | 19.74 | <.001 |
| PSQI 5g | 0.38 | 0.01 | 25.87 | <.001 |
| PSQI 5h | 0.37 | 0.01 | 25.16 | <.001 |
| PSQI 5i | 0.43 | 0.01 | 29.59 | <.001 |
| PSQI 5j | 0.35 | 0.01 | 24.07 | <.001 |
| **DYSFUNCTION** | |  |  |  |
| PSQI 7 | 0.51 | 0.01 | 34.01 | <.001 |
| PSQI 8 | 0.67 | 0.02 | 41.72 | <.001 |
| **MEDICATION** | |  |  |  |
| PSQI 6 | 1 | 0.01 | 107.84 | <.001 |
| **EFFICIENCY** | |  |  |  |
| PSQI 1 | -0.43 | 0.02 | -25.79 | <.001 |
| PSQI 3 | 0.65 | 0.02 | 32.15 | <.001 |
| PSQI 4 | 0.4 |  |  |  |
| **DURATION** | |  |  |  |
| PSQI 4 | 0.79 | 5.66 | 0.14 | .889 |
| **LATENCY** |  |  |  |  |
| PSQI 2 | 0.65 | 0.01 | 46.66 | <.001 |
| PSQI 5a | 0.89 | 0.01 | 61.01 | <.001 |
| **QUALITY** |  |  |  |  |
| PSQI 9 | 1 | 0.01 | 107.84 | <.001 |
| **ESS** |  |  |  |  |
| ESS 1 | 0.75 | 0.01 | 62.27 | <.001 |
| ESS 2 | 0.68 | 0.01 | 54.08 | <.001 |
| ESS 3 | 0.66 | 0.01 | 52.8 | <.001 |
| ESS 4 | 0.58 | 0.01 | 44.68 | <.001 |
| ESS 5 | 0.59 | 0.01 | 45.22 | <.001 |
| ESS 6 | 0.48 | 0.01 | 35.88 | <.001 |
| ESS 7 | 0.67 | 0.01 | 53.86 | <.001 |
| ESS 8 | 0.39 | 0.01 | 28.44 | <.001 |
| **MEQ** |  |  |  |  |
| MEQ 1 | 0.72 | 0.01 | 57.54 | <.001 |
| MEQ 2 | 0.4 | 0.01 | 28.4 | <.001 |
| MEQ 3 | 0.56 | 0.01 | 41.97 | <.001 |
| MEQ 4 | 0.56 | 0.01 | 42.13 | <.001 |
| MEQ 5 | 0.79 | 0.01 | 64.31 | <.001 |

### Appendix G: Regression Coefficients, Associated *p*-Values and Effect Sizes for COSMOS And Established Scales’ Latent Variables: Multivariate Models

|  | COSMOS | | | | Established | | | |
| --- | --- | --- | --- | --- | --- | --- | --- | --- |
| Scale |  | coef | *p* | *η^2^* |  | coef | *p* | *η^2^* |
| Depression | Disturbance | 0.01 | .969 | 0.55 | Disturbance | -0.37 | .620 | 0.62 |
|  | Dysfunction | 1.98 | <.001 | 0.56 | Dysfunction | 5.98 | .390 | 0.39 |
|  | Disorders | 0.24 | .002 | 0.00 | Medication | -0.07 | <.001 | 0.00 |
|  | Bedtime Habits | 0.37 | <.001 | 0.02 | Efficiency | 0.68 | <.001 | 0.00 |
|  | Medication | 0.13 | .047 | 0.00 | Duration | -1.05 | <.001 | 0.00 |
|  | Latency | 0.78 | <.001 | 0.04 | Latency | -0.43 | .021 | 0.02 |
|  | Duration | 0.08 | .003 | 0.00 | Quality | -0.95 | .001 | 0.00 |
|  | Sleep Onset | -0.06 | .284 | 0.00 | ESS | -2.23 | .093 | 0.09 |
|  | Social Jetlag | 0.11 | .040 | 0.00 | MEQ | 1.73 | .013 | 0.01 |
|  | Chronotype | 0.22 | .152 | 0.00 |  |  |  |  |
|  | Quality | 0.61 | <.001 | 0.00 |  |  |  |  |
| Anxiety (GAD-7) | Disturbance | 0.05 | .844 | 0.29 | Disturbance | -0.22 | .285 | 0.29 |
|  | Dysfunction | 1.90 | <.001 | 0.25 | Dysfunction | 6.73 | .148 | 0.15 |
|  | Disorders | 0.94 | <.001 | 0.02 | Medication | -0.21 | <.001 | 0.00 |
|  | Bedtime Habits | 0.87 | <.001 | 0.04 | Efficiency | -0.10 | .001 | 0.00 |
|  | Medication | 0.03 | .775 | 0.00 | Duration | -0.51 | .002 | 0.00 |
|  | Latency | 1.05 | <.001 | 0.01 | Latency | -0.05 | .021 | 0.02 |
|  | Duration | 0.22 | <.001 | 0.00 | Quality | -1.09 | <.001 | 0.00 |
|  | Sleep Onset | -0.25 | .013 | 0.00 | ESS | -2.47 | .046 | 0.05 |
|  | Social Jetlag | 0.54 | <.001 | 0.00 | MEQ | 1.20 | .002 | 0.00 |
|  | Chronotype | 1.01 | <.001 | 0.00 |  |  |  |  |
|  | Quality | 0.73 | .006 | 0.00 |  |  |  |  |
| Compulsive Soothing (CHI-T Factor 2) | Disturbance | -0.09 | .022 | 0.03 | Disturbance | -0.12 | .047 | 0.05 |
|  | Dysfunction | 0.20 | <.001 | 0.13 | Dysfunction | 0.67 | .103 | 0.10 |
|  | Disorders | 0.08 | <.001 | 0.00 | Medication | -0.02 | <.001 | 0.00 |
|  | Bedtime Habits | 0.15 | <.001 | 0.03 | Efficiency | -0.08 | <.001 | 0.00 |
|  | Medication | 0.05 | .008 | 0.00 | Duration | -0.03 | <.001 | 0.00 |
|  | Latency | 0.02 | .469 | 0.00 | Latency | -0.02 | .002 | 0.00 |
|  | Duration | 0.01 | .402 | 0.00 | Quality | -0.10 | <.001 | 0.00 |
|  | Sleep Onset | -0.01 | .735 | 0.00 | ESS | -0.18 | .012 | 0.01 |
|  | Social Jetlag | 0.05 | <.001 | 0.00 | MEQ | -0.03 | <.001 | 0.00 |
|  | Chronotype | -0.02 | .591 | 0.00 |  |  |  |  |
|  | Quality | 0.07 | .119 | 0.00 |  |  |  |  |
| Neuroticism (BIG-5 Factor 3) | Disturbance | 0.03 | .472 | 0.11 | Disturbance | 0.02 | .123 | 0.12 |
|  | Dysfunction | 0.18 | <.001 | 0.08 | Dysfunction | 0.65 | .050 | 0.05 |
|  | Disorders | 0.10 | <.001 | 0.00 | Medication | 0.01 | .001 | 0.00 |
|  | Bedtime Habits | 0.04 | .004 | 0.00 | Efficiency | 0.06 | <.001 | 0.00 |
|  | Medication | 0.01 | .530 | 0.00 | Duration | -0.04 | .002 | 0.00 |
|  | Latency | 0.06 | .019 | 0.01 | Latency | 0.00 | .006 | 0.01 |
|  | Duration | 0.04 | <.001 | 0.00 | Quality | -0.09 | <.001 | 0.00 |
|  | Sleep Onset | 0.03 | .083 | 0.00 | ESS | -0.23 | .011 | 0.01 |
|  | Social Jetlag | 0.02 | .290 | 0.00 | MEQ | 0.17 | .001 | 0.00 |
|  | Chronotype | -0.04 | .385 | 0.00 |  |  |  |  |
|  | Quality | 0.09 | .058 | 0.00 |  |  |  |  |

### Appendix H: Regression Coefficients, Associated *p*-Values and R-Squared for COSMOS And Established Scales’ Latent Variables: Univariate Models

|  | COSMOS | | | | Established | | | |
| --- | --- | --- | --- | --- | --- | --- | --- | --- |
| Scale |  | *R^2^* | coef | *p* |  | *R^2^* | coef | *p* |
| Depression | Disturbance | 0.25 | 2.12 | <.001 | Disturbance | 0.29 | 2.45 | <.001 |
|  | Dysfunction | 0.48 | 3.05 | <.001 | Dysfunction | 0.47 | 3.19 | <.001 |
|  | Disorders | 0.29 | 2.59 | <.001 | Medication | 0.03 | 0.71 | <.001 |
|  | Bedtime Habits | 0.11 | 1.42 | <.001 | Efficiency | 0.03 | 0.79 | <.001 |
|  | Medication | 0.06 | 1.36 | <.001 | Duration | 0.21 | -2.23 | <.001 |
|  | Latency | 0.31 | 2.44 | <.001 | Latency | 0.25 | 2.21 | <.001 |
|  | Duration | 0.02 | -0.39 | <.001 | Quality | 0.27 | 2.09 | <.001 |
|  | Sleep Onset | 0.01 | -0.34 | <.001 | ESS | 0.10 | 1.34 | <.001 |
|  | Social Jetlag | 0.02 | 0.55 | <.001 | MEQ | 0.06 | -1.04 | <.001 |
|  | Chronotype | 0.16 | -2.06 | <.001 |  |  |  | <.001 |
|  | Quality | 0.34 | 2.51 | <.001 |  |  |  | <.001 |
| Anxiety (GAD-7) | Disturbance | 0.18 | 2.60 | <.001 | Disturbance | 0.19 | 2.89 | <.001 |
|  | Dysfunction | 0.31 | 3.56 | <.001 | Dysfunction | 0.28 | 3.61 | <.001 |
|  | Disorders | 0.24 | 3.44 | <.001 | Medication | 0.01 | 0.72 | <.001 |
|  | Bedtime Habits | 0.11 | 2.05 | <.001 | Efficiency | 0.01 | 0.63 | <.001 |
|  | Medication | 0.04 | 1.62 | <.001 | Duration | 0.11 | -2.35 | <.001 |
|  | Latency | 0.20 | 2.87 | <.001 | Latency | 0.17 | 2.64 | <.001 |
|  | Duration | 0.01 | -0.32 | <.001 | Quality | 0.16 | 2.33 | <.001 |
|  | Sleep Onset | 0.00 | -0.26 | <.001 | ESS | 0.05 | 1.46 | <.001 |
|  | Social Jetlag | 0.02 | 0.94 | <.001 | MEQ | 0.02 | -0.96 | <.001 |
|  | Chronotype | 0.08 | -2.08 | <.001 |  |  |  | <.001 |
|  | Quality | 0.20 | 2.81 | <.001 |  |  |  | <.001 |
| Compulsive Soothing (CHI-T Factor 2) | Disturbance | 0.03 | 0.15 | <.001 | Disturbance | 0.04 | 0.19 | <.001 |
|  | Dysfunction | 0.13 | 0.34 | <.001 | Dysfunction | 0.12 | 0.35 | <.001 |
|  | Disorders | 0.07 | 0.27 | <.001 | Medication | 0.00 | 0.05 | <.001 |
|  | Bedtime Habits | 0.09 | 0.27 | <.001 | Efficiency | 0.02 | 0.14 | <.001 |
|  | Medication | 0.01 | 0.14 | <.001 | Duration | 0.05 | -0.22 | <.001 |
|  | Latency | 0.05 | 0.22 | <.001 | Latency | 0.05 | 0.20 | <.001 |
|  | Duration | 0.00 | -0.03 | <.001 | Quality | 0.05 | 0.18 | <.001 |
|  | Sleep Onset | 0.01 | -0.07 | <.001 | ESS | 0.04 | 0.18 | <.001 |
|  | Social Jetlag | 0.02 | 0.15 | <.001 | MEQ | 0.03 | -0.17 | <.001 |
|  | Chronotype | 0.06 | -0.27 | <.001 |  |  |  | <.001 |
|  | Quality | 0.06 | 0.22 | <.001 |  |  |  | <.001 |
| Neuroticism (BIG-5 Factor 3) | Disturbance | 0.09 | 0.29 | <.001 | Disturbance | 0.10 | 0.33 | <.001 |
|  | Dysfunction | 0.14 | 0.37 | <.001 | Dysfunction | 0.14 | 0.39 | <.001 |
|  | Disorders | 0.11 | 0.36 | <.001 | Medication | 0.02 | 0.11 | <.001 |
|  | Bedtime Habits | 0.03 | 0.17 | <.001 | Efficiency | 0.00 | 0.08 | <.001 |
|  | Medication | 0.02 | 0.19 | <.001 | Duration | 0.05 | -0.25 | <.001 |
|  | Latency | 0.10 | 0.31 | <.001 | Latency | 0.09 | 0.29 | <.001 |
|  | Duration | 0.00 | -0.03 | 0.001 | Quality | 0.08 | 0.26 | <.001 |
|  | Sleep Onset | 0.00 | -0.02 | 0.167 | ESS | 0.03 | 0.16 | <.001 |
|  | Social Jetlag | 0.01 | 0.08 | <.001 | MEQ | 0.01 | -0.10 | <.001 |
|  | Chronotype | 0.05 | -0.26 | <.001 |  |  |  |  |
|  | Quality | 0.12 | 0.33 | <.001 |  |  |  |  |

**Figure 1**

*Effect sizes of regression coefficients for COSMOS and established scales’ subscales when estimating psychological symptoms and traits using independent regression models for each subscale.*

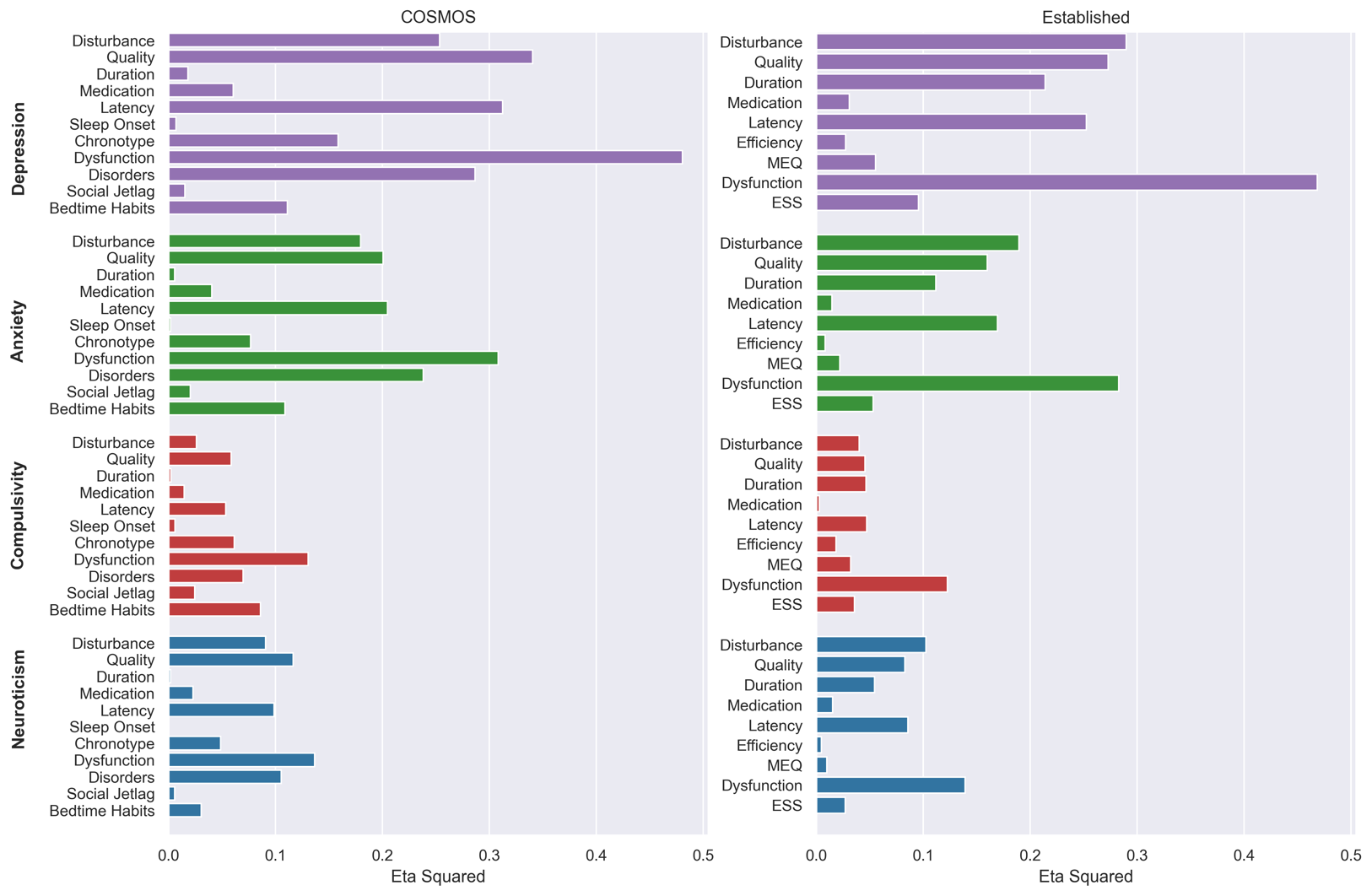
